# A High-Throughput Broad Neutralizing Antibody Assay for Detecting SARS-CoV-2 Variant Immunity in Population

**DOI:** 10.1101/2023.08.07.23293304

**Authors:** Xiaohan Zhang, Yajie Wang, Mansheng Li, Haolong Li, Xiaomei Zhang, Xingming Xu, Qingqing Ma, Di Hu, Yan Jia, Te Liang, Yunping Zhu, Surbhi Bihani, Sanjeeva Srivastava, Manuel Fuentes, Yongzhe Li, Xiaoxu Han, Bingwei Wang, Xiaobo Yu

**Affiliations:** State Key Laboratory of Medical Proteomics, Beijing Proteome Research Center, National Center for Protein Sciences-Beijing (PHOENIX Center), Beijing Institute of Lifeomics, Beijing, 102206, China; School of Medicine, Nanjing University of Chinese Medicine, Nanjing 210023, China; Department of Clinical Laboratory, Beijing Ditan Hospital, Capital Medical University, Beijing 100015, China; Department of Clinical Laboratory, Peking Union Medical College Hospital, Chinese Academy of Medical Science & Peking Union Medical College, Beijing 100730, China; State Key Laboratory for Diagnosis and Treatment of Infectious Diseases, National Clinical Research Center for Laboratory Medicine, The First Hospital of China Medical University, China Medical University, Shenyang, 110001, China; ProteomicsEra Medical Co., Ltd., Beijing, 102206, China; Department of Biosciences and Bioengineering, Indian Institute of Technology Bombay, Mumbai, Maharashtra, 400076, India; Department of Medicine and General Cytometry Service-Nucleus, CIBERONC ISCIII, Cancer Research Center (IBMCC/CSIC/USAL/IBSAL), Salamanca, 37007, Spain; School of Basic Medicine Sciences, Anhui Medical University, Hefei, Anhui 230031, China; College of Life Sciences, Hebei University, Baoding, 071002, China; School of Basic Medicine, Qingdao University, Qingdao 266071, China

**Keywords:** COVID-19, SARS-CoV-2, flow cytometry, neutralizing antibody, variant

## Abstract

Detecting neutralizing antibodies (NAbs) to SARS-CoV-2 variants that are evolved is crucial to know the escape of host immunity to the newly arising variants. To address this need, we developed a high-throughput broad neutralizing antibody (bNAb) assay using flow cytometry with magnetic-fluorescent microspheres for detecting NAbs against diverse SARS-CoV-2 variants. The assay is rapid, reliable, 35-fold more sensitive than Luminex technology. Our results highly correlated with IgG serological assay (R = 0.90) , the FDA-approved cPass sVNT assay (R = 0.92), pseudovirus-based neutralizing assay (R = 0.96, 0.66, 0.65) and live virus based neutralization assay (R = 0.79, 0.64). When applied to 56 healthy individuals receiving third-dose vaccines (18 CoronaVac; 38 ZF2001) and 35 HIV patients with breakthrough infection of COVID-19 (16 with CD4+ < 350 cells/µL; 19 with CD4+ > 500 cells/µL), results showed that the Omicron BA.1-BA.5 variants exhibited significant resistance to inactivated vaccines in healthy individuals. In HIV patients, the breakthrough infection of Omicron BA.5.2 or BF.7 variants can induce broad neutralizing activity to non-Omicron and Omicron variants together with vaccination. Notably, the levels of NAbs against most of SARS-CoV-2 variants are much lower in the decreased immunity of HIV patients (CD4+ < 350 cells/µL) compared to the recovered immunity (CD4+ > 500 cells/µL), indicating that maintenance of the immune system is crucial for NAb production. Altogether, our high-throughput proteomics platform represents a powerful tool for the detection of bNAbs in the population and may inform the development of more effective COVID-19 vaccines and vaccination strategies in the future.

## INTRODUCTION

The emergence of severe acute respiratory syndrome coronavirus 2 (SARS-CoV-2) and the ensuing coronavirus 2019 (COVID-19) pandemic have profoundly impacted global health^1^. Although the acute crisis phase has subsided, persistent viral transmission and continuous SARS-CoV-2 evolution remain significant challenges as evidenced by emerging variants including EG.5.1, JN.1, and KP.2 that continue to threaten public health^2^. According to reports, 17.13% (95%CI, 7.55–26.71%) of asymptomatic infected individuals may have long-term health consequences^3^, which may include symptoms such as fatigue, brain fog, dizziness, gastrointestinal symptoms, as well as others^4^. Therefore, it would remain significant to judge the ability of newly arising variants to escape the immunity in population defensed by previous vaccination and/or infections^5^.

Neutralizing antibodies (NAbs) of SARS-CoV-2 are a subset of antibodies that block viral entry by inhibiting the interaction between the SARS-CoV-2’s surface protein, Spike, and the host cell’s receptor, angiotensin-converting enzyme 2 (ACE2). Such NAbs can be produced following COVID-19 vaccination or SARS-CoV-2 infection. Identifying SAR-CoV-2 variant specific NAbs is critical for assessing vaccine efficacy, evaluating individual and population immunity, monitoring variant susceptibility, and information public health measures to control the spread of COVID-19^6,7^. However, traditional methods of NAb detection, such those that use a live virus or a pseudovirus-based neutralization assay, are slow, expensive, carry a potential risk of infection, and may require specialized biosafety level 3 (BSL3) facilities^8–11^.

As an alternative approach, enzyme-linked immunosorbent assay (ELISA) can perform the *in vitro* neutralizing testing using purified Spike, ACE2, or Spike’s receptor-binding domain (RBD) that interacts directly with ACE2 during viral entry. Several commercial ELISA-based surrogate virus neutralization tests (sVNT) have been developed, including TECO sVNT (TECO Medical) and cPass sVNT (GenScript), which have been approved by the United States’ (U.S.) Food and Drug

Administration (FDA) or received Conformite Europeenne (CE) marking for use in the European Union^12,13^. Unfortunately, ELISA-based methods only detect a single variant at a time, making them a time-consuming option when multiple SARS-CoV-2 variants are analyzed^9^.

To address this concern, Fenwick *et al.* developed a multiplexed *in-vitro* quantitative neutralization assay using Luminex technology in 2021. The assay enables the detection of NAbs targeting various Spike mutations, including D614G, D614G plus M153T, N439K, S477N, S477R, E484K, S459Y, N501T, N501Y, K417N, 60-70, P681H, Y453F, and their combinations. The results obtained from this assay correlated with pseudovirus neutralization (R^2^=0.65) and live virus infection (R^2^ = 0.825) assays^10^.

In a similar vein, Lynch et al. developed a multiplexed surrogate virus neutralization test (plex-sVNT) for detecting NAbs to seven SARS-CoV-2 variants, namely, wild type (WT), Alpha, Beta, Gamma, Delta, Kappa, and Epsilon. The results highly correlated (> 96%) with those obtained from a plaque reduction neutralization test (PRNT)^11^.

However, it should be noted that these technologies have limitations as they require access to specific Luminex and Bio-Rad instruments. This requirement restricts their adoption primarily to specialized laboratories equipped with these systems, limiting broader accessibility and high-throughput screening potential. Moreover, some recent Omicron variants have not been accessible for analysis using these methods to date^14^.

To overcome these limitations, specifically the need for a multiplexed, high-throughput assay that is accessible to a wider range of laboratories using standard equipment and capable of covering emerging variants, we developed a high-throughput broad NAb (bNAb) assay. This approach leverages the widespread availability and standardization of flow cytometers in research and clinical settings. Our assay enables the systematic and simultaneous detection of NAbs to eleven SARS-CoV-2 variants of concern (VOCs), including D614G, Alpha, Beta, Gamma, Delta, Kappa, Omicron BA.1-BA.5, in a format designed to be reliable, cost-effective, sensitive, and crucially, utilizable with a standard flow cytometer^15^. This addresses the core accessibility barrier of the Luminex-based methods.

We conducted a comparative analysis of NAb detection using our SARS-CoV-2 bNAb assay with commercial ELISA-based serology, U.S. FDA-approved cPass sVNT, pseudovirus-based neutralization and live virus based neutralization assays. This validation demonstrates the performance and reliability of our flow cytometry platform as a robust alternative or complement to existing technologies. Furthermore, we explored the applications of the SARS-CoV-2 bNAb assay for screening therapeutic antibodies, assessing their neutralizing activity SARS-CoV-2. Additionally, we analyzed the broad responses of serological NAbs to SARS-CoV-2 variants in individuals who received the third dose of either the inactivated vaccine (Sinovac-CoronaVac) or the recombinant subunit vaccine (ZF2001) for COVID-19. Finally, we assessed the serum NAb inhibition rates (%) against SARS-CoV-2 variants in the decreased immunity (CD4+ < 350 cells/µL) and recovered immunity (CD4+ > 500 cells/µL) of HIV patients with breakthrough infection of COVID-19 (Figure S1).

## RESULTS

### Schematic illustration of SARS-CoV-2 bNAb assay

In the SARS-COV-2 bNAb assay, illustrated schematically in Figure 1A, the trimerized Spike proteins of six non-Omicron variants (D614G, Alpha, Beta, Kappa, Gamma, Delta) and five Omicron variants (BA.1, BA.2, BA.3, BA.4, BA.5) were expressed from human embryonic kidney 293 (HEK293) cells, purified, and coupled to the magnetic-fluorescent beads, as previously reported^10,16^. During the NAbs detection process, all coupled beads were mixed together and incubated with samples containing NAbs. The NAbs bound to the Spike trimer proteins, preventing their interaction with the biotinylated ACE2 receptors, which were coupled to streptavidin-phycoerythrin (SA-PE). The inhibition rate (%) was calculated as follows: Inhibition rate (%) = (1 - signal of NAb inhibition on Spike-ACE2 binding / signal of Spike-ACE2 binding) × 100%.

**Figure 1.**
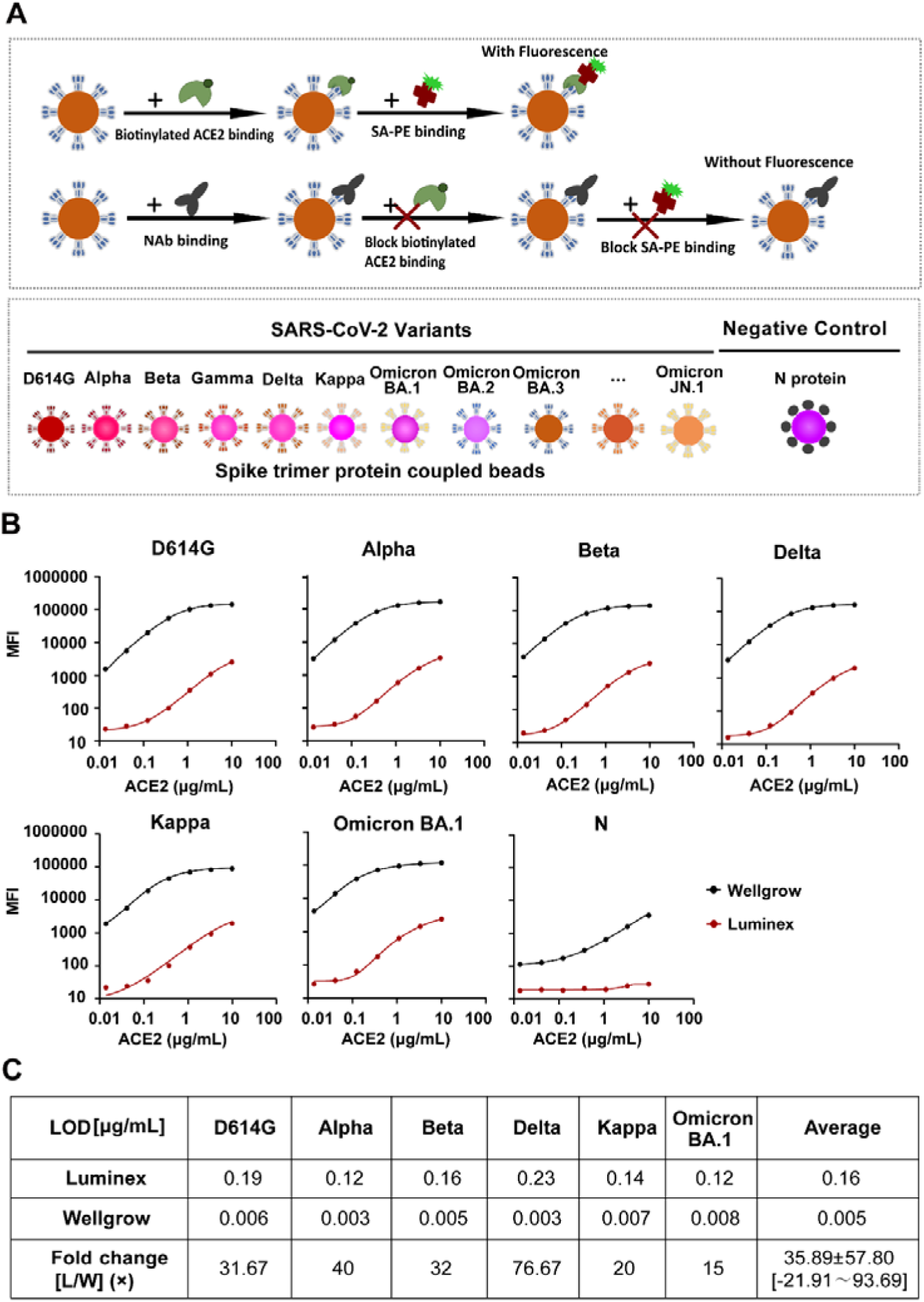
Schematic illustration of the high-throughput SARS-CoV-2 bNAb assay. (A) Workflow of the SARS-CoV-2 bNAb assay. The Spike-ACE2 interaction is a direct binding reaction between Spike trimer proteins and biotinylated ACE2. Upon the addition of the NAbs, the beads coupled with SARS-CoV-2 variants Spike trimer proteins are incubated with NAbs. The NAbs with neutralizing activity will block the Spike interaction with biotinylated ACE2 and SA-PE. The signal intensities of the Spike trimer-ACE2 interaction are inversely proportional to the level of NAbs. During the NAbs detection process, all coupled beads were mixed together and incubated with samples containing NAbs. The NAbs bound to the Spike trimer proteins, preventing their interaction with the biotinylated ACE2 receptors, which were coupled to SA-PE. (B) Dose relationship curve of Spike-ACE2 interactions using Wellgrow and Luminex platforms. The x-axis represents the concentrations of ACE2. The y-axis represents the MFI of the SARS-CoV-2 Spike trimer-ACE2 interaction. (C) A comparison of the sensitivity to detect Spike-ACE2 interactions with the Wellgrow and Luminex platforms. N protein, nucleocapsid protein. bNAb, broad neutralizing antibody. ACE2, angiotensin-converting enzyme 2. SA-PE, streptavidin-phycoerythrin. MFI, median fluorescence intensity. LOD, lowest detection limit.

The sensitive detection of Spike-ACE2 interactions is essential for assessing the presence of NAbs in clinical samples. To address this requirement, we coupled the Spike protein from six SARS-CoV-2 variants (D614G, Alpha, Beta, Kappa, Delta, Omicron BA.1) onto magnetic-fluorescent beads from Luminex and Shenzhen Wellgrow Technology Co., Ltd. (“Wellgrow”), and then incubated with different concentrations of ACE2. Magnetic-fluorescent beads with the SARS-CoV-2 nucleocapsid (N) protein, rather than the Spike protein, was employed as the negative control. The binding signals were measured using Luminex-200 and Wellgrow flow cytometry. Notably, the fluorescent signal intensity and signal to noise ratio (SNR) were much higher with the Wellgrow platform than the Luminex platform when ACE2 concentrations ranged from 0.01 µg/mL to 100 µg/mL (Figure 1B, Figure S2 and Table S6).

Furthermore, we calculated the sensitivity or the lowest detection limit (LOD) of each method, in which the LOD was equal to the signal of the buffer control plus ten standard deviations (Figure 1C). The data showed that the average LOD of our assay using the Wellgrow platform was 35.89 ± 57.8 [-21.91 ∼ 93.69]-fold higher than the Luminex platform for all variants (D614G, Alpha, Beta, Kappa, Delta, Omicron BA.1).

At last, we compared the difference of NAb titers obtained by detecting the inhibition of antibody #26 to Spike-ACE2 interaction. The results showed that the IC50 obtained by two platforms are highly consistence with the r correlation of 0.9988 (Figure S2D). Therefore, we chose the Wellgrow platform as the proteomics platform for the SARS-CoV-2 bNAb assay in this study.

### Mapping Spike-ACE2 interactions of SARS-CoV-2 variants

Using our platform, we conducted a systematic investigation of the interactions between ACE2 and Spike trimer proteins from eleven SARS-CoV-2 variants. This was achieved by incubating a bead array with different concentrations of ACE2 (Figure 2). The results demonstrated that the fluorescent signals from the Spike trimer proteins increased with increasing concentrations of ACE2, indicating the formation of Spike-ACE2 complexes on the beads (Figure 2A).

**Figure 2.**
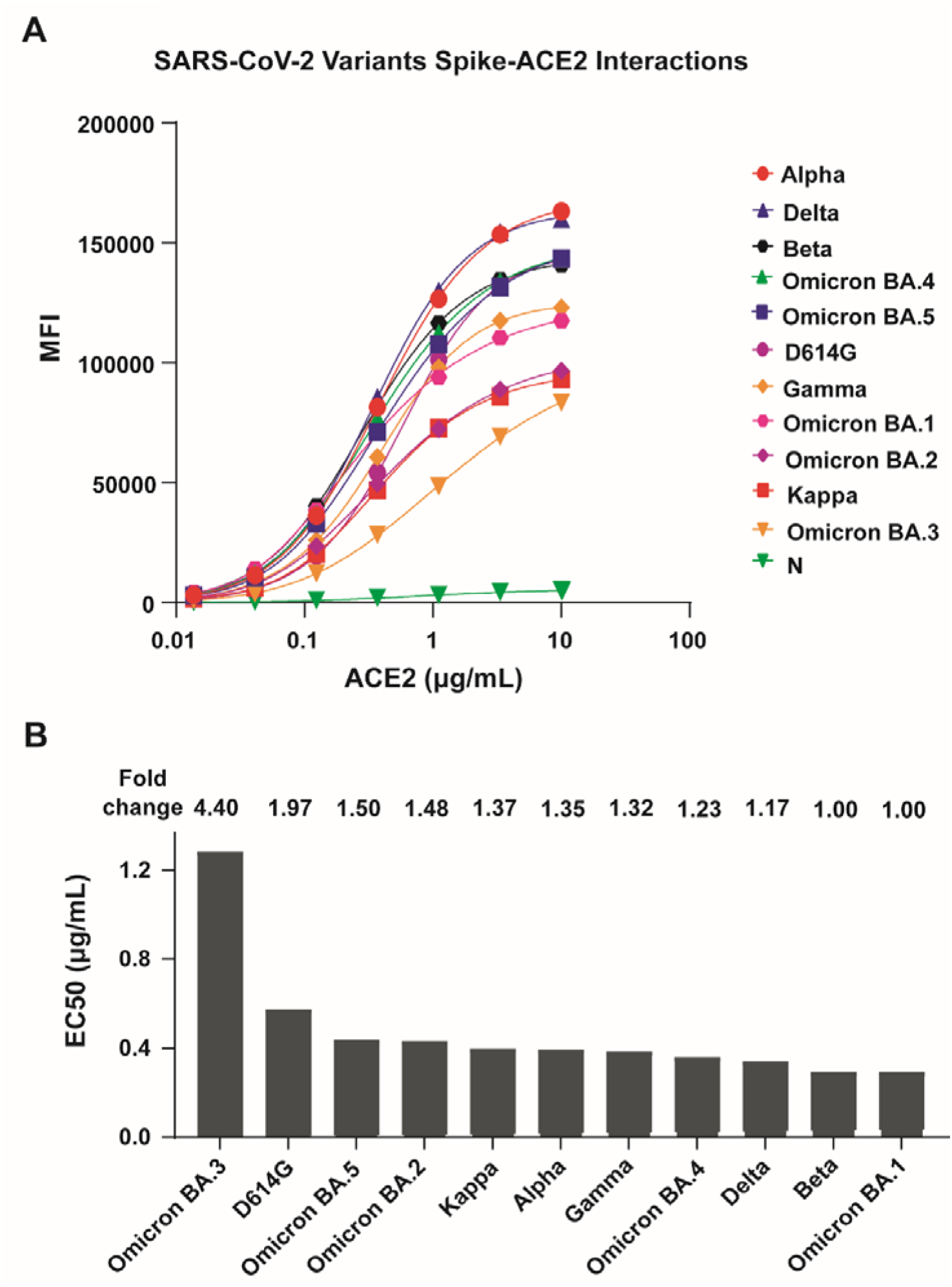
Spike trimer-ACE2 interactions across different SARS-CoV-2 variants and ACE2 concentrations. (A) Dose relationship curve of the interactions between ACE2 and six SARS-CoV-2 non-Omicron variants and five Omicron variants. The x-axis represents the concentrations of the host ACE2 receptor. The y-axis represents the MFI of the SARS-CoV-2 Spike trimer-ACE2 interaction. (B) The level of binding Affinity of SARS-CoV-2 variant Spike-ACE2 interactions. The x-axis represents the SARS-CoV-2 variants. The y-axis represents the EC50 calculated from SARS-CoV-2 variant Spike-ACE2 interactions. N protein, nucleocapsid protein. angiotensin-converting enzyme 2. MFI, median fluorescence intensity. EC50, half-maximal effective concentration.

Spike mutations were found to influence the interaction dynamics. At the maximal concentration of 10 µg/mL ACE2, Alpha and Delta variants exhibited the highest signals, followed by Beta, BA.4, BA.5, D614G, Gamma, BA.1, BA.2, and Kappa, with BA.3 showing the lowest signal. In contrast, the N protein, used as a negative control, consistently displayed the lowest signal regardless of ACE2 concentration.

To further assess the binding affinity, we calculated the half-maximal effective concentration (EC50) using the standard curve for each SARS-CoV-2 variant and expressed values relative to Omicron BA.1 (1.00×) (Figure 2B). The results showed that Omicron BA.1 has the highest binding affinity with an EC50 of 0.2913 µg/mL, followed by Beta (0.2915 µg/mL, 1.00×), Delta (0.3415 µg/mL, 1.17×), Omicron BA.4 (0.3592 µg/mL, 1.23×), Gamma (0.3833 µg/mL, 1.32×), Alpha (0.3940 µg/mL, 1.35×), Kappa (0.3983 µg/mL, 1.37×), Omicron BA.2 (0.4307 µg/mL, 1.48×), Omicron BA.5 (0.4359 µg/mL, 1.50×), D614G (0.5729 µg/mL, 1.97×), and Omicron BA.3 (1.2830 µg/mL, 4.40×). These datas demonstrate significant quantitative differences in the binding affinity of Spike-ACE2 interactions across the tested SARS-CoV-2 variants. The results are in accordance with surface plasmon resonance data in which BA.1 has the strongest interaction with ACE2 with a dissociation constant (K_D_) of 2.10 nM, followed by BA.2 (2.21 nM) and D614G (5.20 nM)^17^. Similar results were obtained by Mahalingam et al., who used ELISA to determine the EC50 of ACE2 to Spike Omicron (0.38 nM), Delta (0.48 nM), and WT (2.28 nM)^18^. Collectively, our data provide a comprehensive landscape of Spike trimer-ACE2 interactions for SARS-CoV-2 VOCs, as classified by the U.S. Centers for Disease Control and Prevention (CDC) (https://www.cdc.gov/coronavirus/2019-ncov/variants/variant-classifications.html). It should be noted that the Epsilon variant was not included in this work due to the unavailability of purified Spike trimer protein at the time of executing experiments.

In addition, to demonstrate the specificity of our assay, we tested the binding ability of ACE2 to the Spike proteins from MERS, SARS-CoV and six SARS-CoV-2 variants. The results showed that the binding signal of ACE2 to six SARS-CoV-2 variant Spike proteins was much higher than that of SARS-CoV and MERS. The binding signal of SARS-CoV was 3.38-5.74 times lower than that of SARS-CoV-2 variants, due to the homology of Spike sequence. There was no binding signal observed between ACE2 and MERS, similar to the negative control, N protein (Figure S3). All these results are in accordance with the reported results^19^, and demonstrate our assay is specific.

### Comparison of SARS-CoV-2 bNAb assay to ELISA, cPass sVNT, pseudovirus-based neutralization and live virus based neutralization assays

To assess the reliability of the SARS-CoV-2 bNAb assay, we compared the SARS-CoV-2 bNAb assay to an ELISA that was used to detect anti-Spike IgG antibodies^20^. The results showed a high correlation between the levels of D614G NAbs detected using SARS-CoV-2 bNAb assay and the levels of anti-Spike IgG antibodies obtained by the ELISA (R = 0.90, p < 2.2e-16) in a cohort of 75 individuals who provided clinical serum samples approximately 180-270 days after the third dose of the COVID-19 vaccine (Figure 3A). ROC analysis demonstrated excellent diagnostic performance of the SARS-CoV-2 NAb assay using ELISA as reference standard, yielding an AUC of 0.894 (95% CI: 0.813-0.975) (Figure S4A).

**Figure 3.**
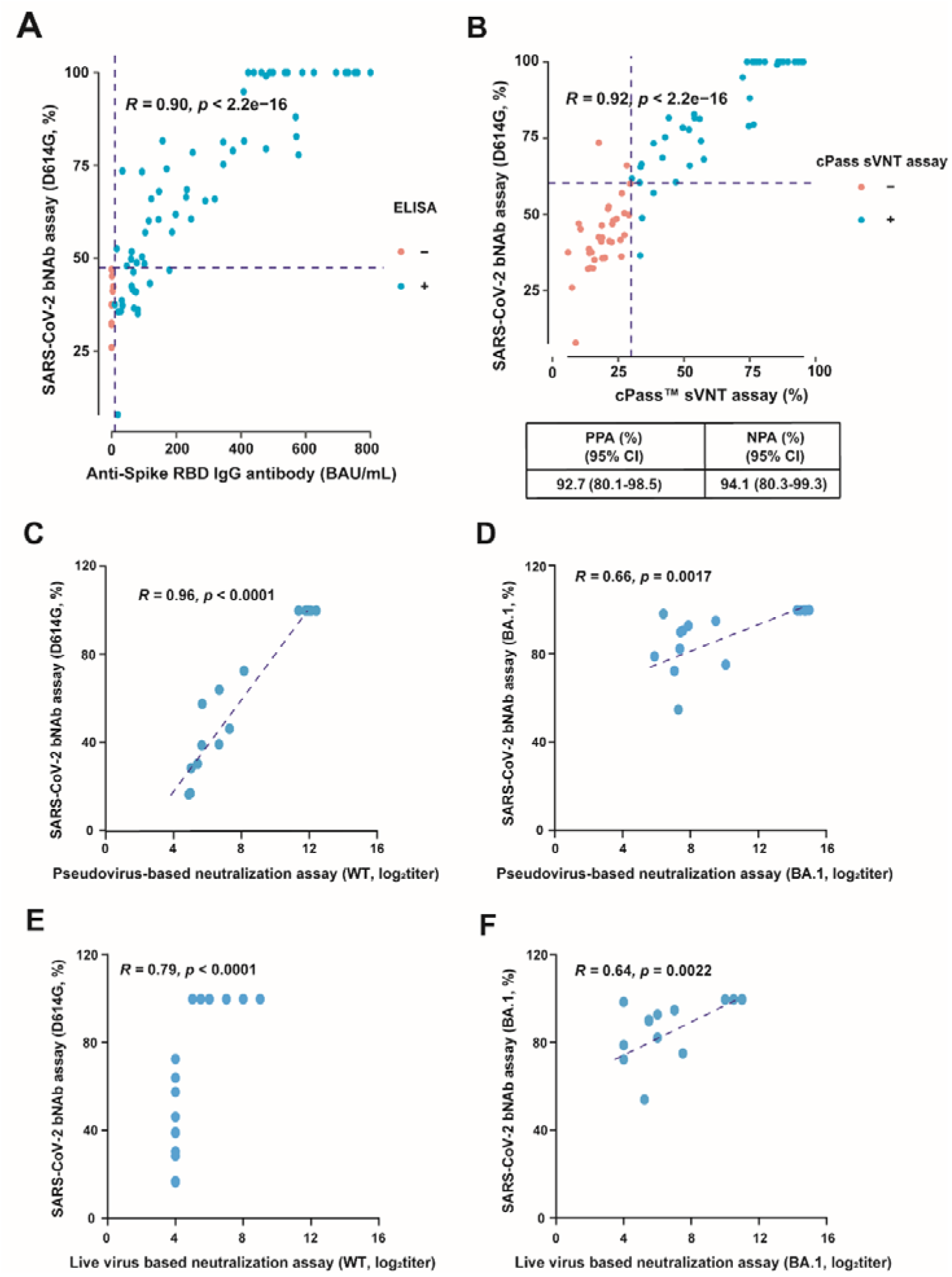
Comparison of SARS-CoV-2 bNAb assay to ELISA, cPass sVNT, pseudovirus-based neutralization and live virus based neutralization assays. (A) Correlation between NAbs inhibition rate (%) from the SARS-CoV-2 bNAb assay and the levels of anti-Spike IgG antibodies obtained by the ELISA in 75 serum samples obtained approximately 180-270 days after the third dose of COVID-19 vaccine. (B) Correlation between the levels of NAbs obtained by SARS-CoV-2 bNAb assay and the FDA-approved cPass sVNT assay in 75 serum samples obtained approximately 180-270 days after the third dose of COVID-19 vaccine. The concordance of NAb detection using the ELISA and sVNT assays was calculated as the PPA and NPA, respectively. The blue and red dots represent positive (neutralizing activity) and negative (non-neutralizing) cases as determined by the comparative analysis, respectively. (C-D) Correlation between NAbs inhibition rate (%) from the SARS-CoV-2 bNAb assay and NAb titers from pseudovirus-based neutralization assays in serum samples from mRNA-vaccinated mice (n = 20). (E-F) Correlation between NAbs inhibition rate (%) from the SARS-CoV-2 bNAb assay and NAb titers from live virus based neutralization assays in serum samples from mRNA-vaccinated mice (n = 20). N protein, nucleocapsid protein. bNAb, broad neutralizing antibody. WT, wild type. RBD, receptor-binding domain. PPA, positive percent agreement. NPA, negative percent agreement.

Next, within the same cohort, we compared the NAb results obtained from the SARS-CoV-2 bNAb assay to those obtained from the FDA-approved cPass sVNT assay^9,12^. The data obtained using the SARS-CoV-2 bNAb assay and cPass sVNT assay demonstrated a higher correlation to each other (R = 0.92, p < 2.2 e-16) compared to the assay using Luminex technology (R = 0.85), as previously reported by Marien et al.^21^. The positive percent agreement (PPA) was 92.7% (80.1% - 98.5%) and the negative percent agreement (NPA) was 94.1% (80.3% - 99.3%) (Figure 3B). ROC analysis confirmed high accuracy of the SARS-CoV-2 NAb assay using cPass sVNT as reference standard, with an AUC of 0.964 (95% CI: 0.923-1.000) (Figure S4B). The findings suggest that some of the anti-Spike IgG antibodies detected by the ELISA may lack neutralizing activity and therefore were not detected by SARS-CoV-2 bNAb assay^22^. Overall, these results demonstrate the reliability of the SARS-CoV-2 bNAb assay as an *in vitro* platform for detecting serum NAbs.

Additionally, we compared the inhibition rate (%) of NAbs obtained by SARS-CoV-2 bNAb assay to the NAb titers obtained by pseudovirus-based neutralization assay or live virus based neutralization assay. For 10 serum samples from mice administered with WT mRNA vaccines and 10 serum samples from mice administered BA.1 mRNA vaccines, a high correlation was found between the D614G NAbs inhibition rate (%) detected using SARS-CoV-2 bNAb assay and WT NAb titers detected by pseudovirus-based neutralization assay (R = 0.96, p < 0.0001) and live virus based neutralization assay (R = 0.79, p < 0.0001) (Figures 3C, 3E). Similarly, notable correlations were observed between the levels of BA.1 NAbs detected using SARS-CoV-2 bNAb assay and pseudovirus-based neutralization (R = 0.66, p = 0.0017) and live virus based neutralization assays (R = 0.64, p = 0.0022) (Figures 3D, 3F). Then, we compared the inhibition rate (%) of NAbs obtained by SARS-CoV-2 bNAb assay with the NAb titers obtained by pseudovirus-based neutralization assay in 135 clinical serum samples collected from individuals 28 days after receiving the second dose of the COVID-19 vaccine. We detected serum NAbs against the D614G variant using both assays. The results showed a correlation coefficient (R) of 0.65, demonstrating the consistency between the results obtained through inhibition and those obtained with NAb titers (Figure S5).

### Identification of broadly neutralizing monoclonal/polyclonal antibodies against SARS-CoV-2 variants

To demonstrate the application of SARS-CoV-2 bNAb assay in identifying monoclonal/polyclonal antibodies for potential treatment purposes, we tested six anti-Spike antibodies that are available in the laboratory^23^ (Figure 4A). The results revealed that three antibodies (#26, #20, #21) have neutralizing activity. Antibodies #20 and #21 inhibited ACE2 binding to D614G, Alpha, Beta, Kappa and Delta variants, but were not effective in inhibiting the ACE2 binding to the Omicron BA.1 variant. Antibody #26 exhibited the ability to inhibit Spike-ACE2 interactions for D614G, Alpha, Beta, Kappa,Delta and Omicron BA.1 variants.

**Figure 4.**
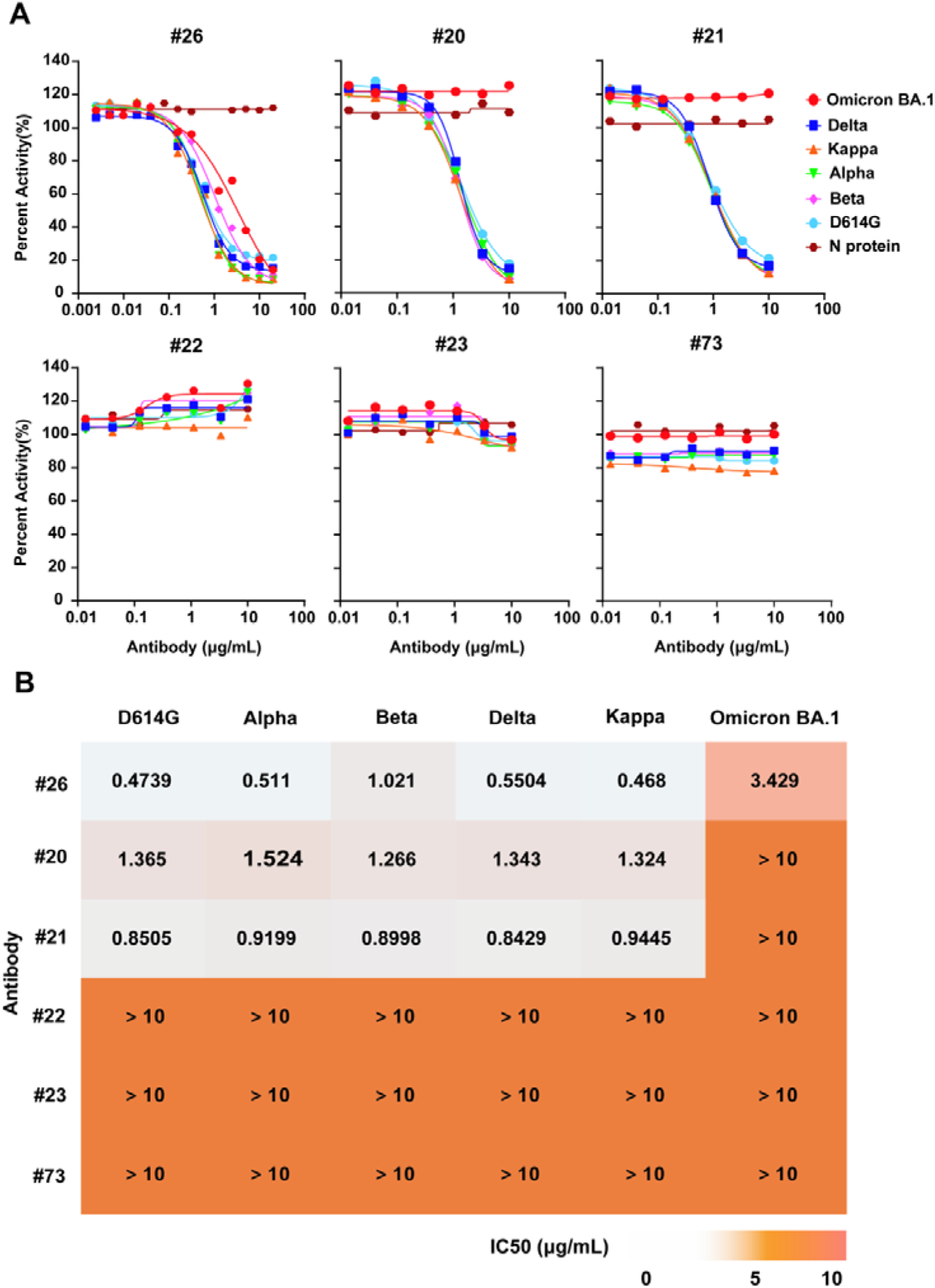
Identification of monoclonal/polyclonal antibodies with broad neutralizing capability. (A) Dose relationship curve demonstrating the ability of the NAb to inhibit SARS-CoV-2 variant Spike ACE2 interactions. The x-axis represents the concentrations of the NAb. The y-axis represents the percent activity (%) of the NAb. (B) Heatmap showing the IC50 of NAbs against different SARS-CoV-2 variants. The detail of monoclonal/polyclonal **antibodies** are provided in Table S5. N protein, nucleocapsid protein. NAb, neutralizing antibody. IC50, half-maximal inhibitory concentration.

Subsequently, the half-maximal inhibitory concentration (IC50) values were calculated and represented in a heatmap (Figure 4B). The results showed that antibody #26 possessed broad neutralizing activity to all tested SARS-CoV-2 variants (Kappa, D614G, Alpha, Delta, Beta, Omicron BA.1). The IC50 to D614G (0.47 µg/mL) was confirmed with ELISA (0.35 µg/mL) and pseudovirus-based neutralization assay (0.21 µg/mL) in our previous work^23^. While the IC50 of antibody #26 to the Omicron variant was ten-fold lower than non-Omicron variants. These findings underscore the ability of our assay to rapidly screen for monoclonal/polyclonal antibodies with potential therapeutic applications.

### Evaluation of serum NAbs in healthy individuals receiving the third dose vaccination and HIV patients with breakthrough infection of COVID-19

We tested the serum NAbs from 56 healthy individuals who received two doses of the inactivated vaccine (Sinovac-CoronaVac) and boosted with a third dose with either an inactivated vaccine or recombinant subunit vaccine (ZF2001) (Table 1)^24^. The inhibition rate (%) was calculated by deducting the background noise using 147 serum samples collected prior to the COVID-19 pandemic^25^. The results showed that the third dose vaccination elicited a strong protective immune response against the non-Omicron variants with the highest inhibition rate (%) of D614G (reference = 1.00×), followed by Gamma, Alpha, Delta, and Kappa variants (all > 0.80×). Omicron variants exhibited substantial immune escape, with BA.4 showing the greatest escape (0.33×), followed by BA.5 (0.35×), BA.1 (0.43×), BA.3 (0.41×), and BA.2 (0.44×), indicating the resistance of these variants to the Sinovac-CoronaVac vaccine and the ZF2001 vaccine (Figure 5A).

**Figure 5.**
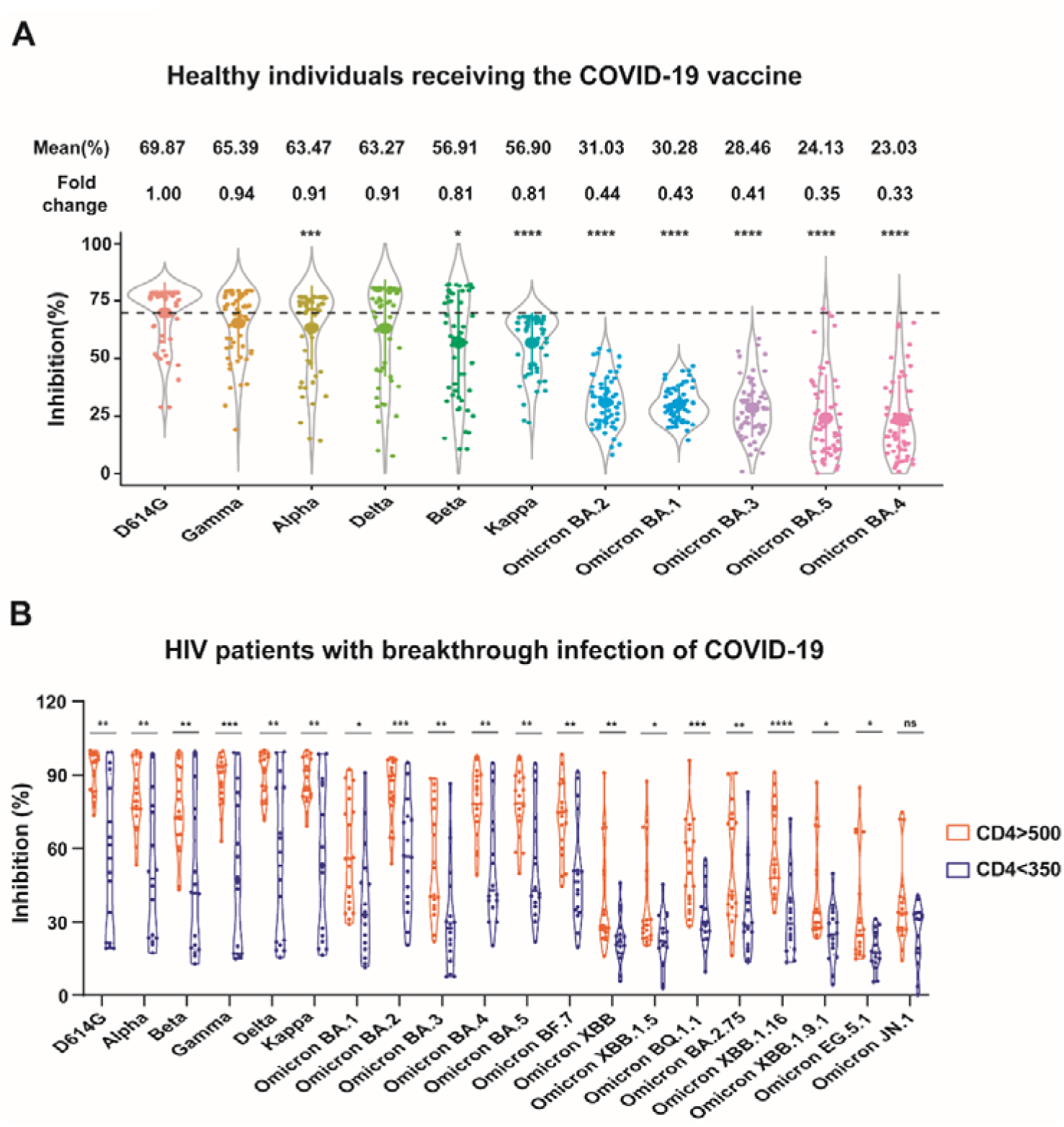
Detection of serum NAbs in vaccinated healthy individuals and HIV patients with breakthrough infection of COVID-19. (A) Boxplot showing the distribution of inhibition rate (%) of serum NAbs against SARS-CoV-2 variants in 56 vaccinated individuals. The significance was performed using the Student’s *t*-test. (B) Boxplot showing the distribution of inhibition rate (%) of serum NAbs against SARS-CoV-2 variants in 35 HIV patients infected with Omicron BA.5.2 and BF.7 variants, incluing 19 immunological responders (CD4+ T cell counts > 500 cells/µL) and 16 poor immunological responders (CD4+ T cell counts < 350 cells/µL). NAb, neutralizing antibody. Asterisks denote statistical differences (*p < 0.05, **p < 0.01, ***p < 0.001, ****p <0.0001, ns: not signiffcant).

**Table 1.**
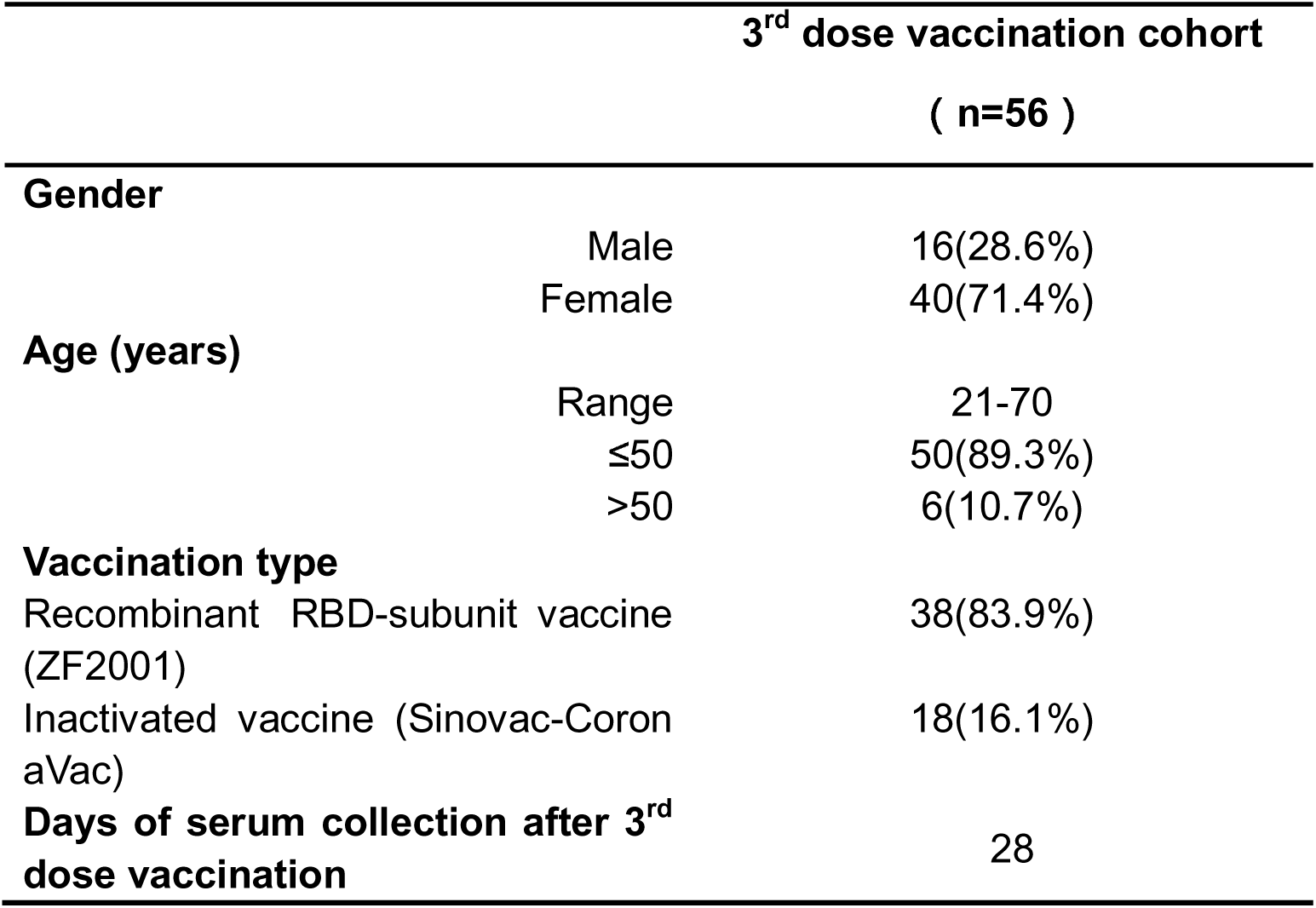
Clinical serum samples of 56 healthy individuals receiving third-dose vaccines employed in this study.

Next, we compared the inhibition rate (%) of serum NAbs in individuals who received either a third dose of inactivated vaccine or recombinant vaccine; no significant difference was observed (Figure S6). These findings highlight the immunological resistance of different SARS-CoV-2 variants to the Sinovac-CoronaVac and ZF2001 vaccines, and suggest the need for developing a new vaccine that provides broad protection against SARS-CoV-2 variants.

Furthermore, in order to demonstrate the specificity of our SARS-CoV-2 bNAb assay, we detected the serum NAbs from the individuals before vaccination (n = 147) and after three-dose vaccination (n = 56). The results showed that the serum NAb levels were significantly higher in the vaccination group than un-vaccination group (p < 0.0001) (Figure S7), indicating our SARS-CoV-2 bNAb assay is highly specific for the detection of serum NAbs.

Finally, we tested the serum NAbs from 35 HIV patients who received three-dose inactivated vaccination and experienced Omicron BA.5.2 or BF.7 breakthrough infections (Table 2)^26^. Subsequently, we assessed the inhibition rate (%) on 20 SARS-CoV-2 variants, which included Omicron EG.5.1 and JN.1. The results showed that Omicron EG.5.1 and JN.1 exhibited strong immune escape with low inhibition rates. Notably, individuals with recovered immunity (CD4+ > 500 cells/µL) exhibited significantly higher serum NAb inhibition rates against most of SARS-CoV-2 variants compared to individuals with decreased immunity (CD4+ < 350 cells/µL) (p < 0.05) (Figure 5B). This demonstrates that while breakthrough infection enhances variant coverage, optimal immune reconstitution, rather than mere infection exposure, is essential for generating cross-protective immunity against highly evasive variants like EG.5.1 and JN.1 in HIV populations.

**Table 2.**
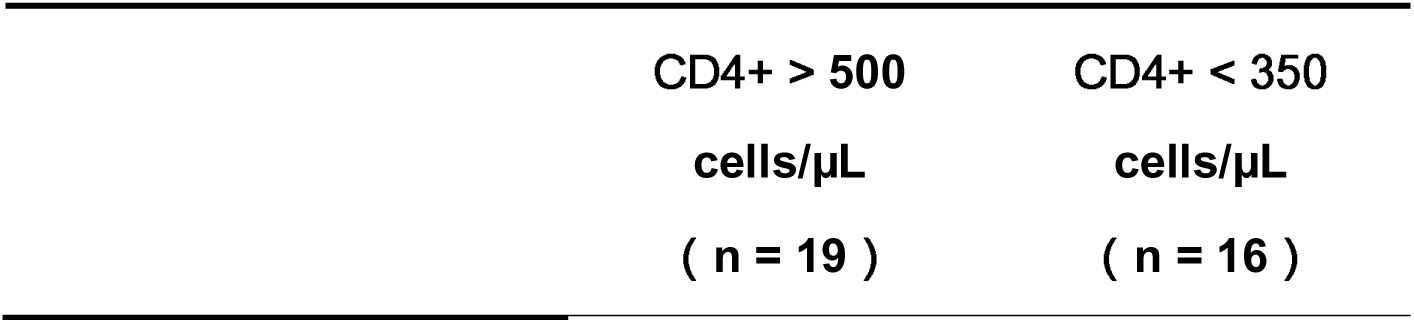
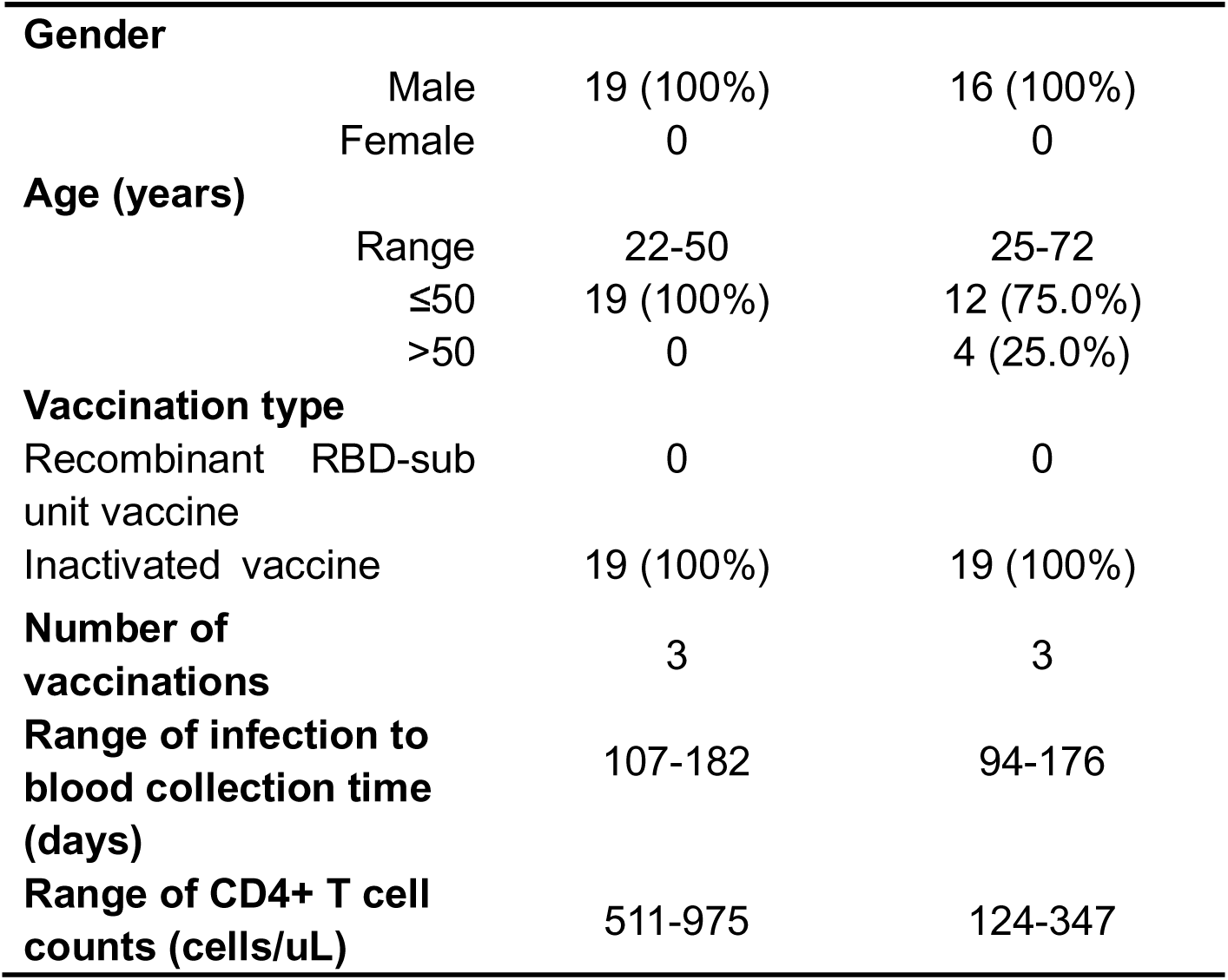
Clinical serum samples of 35 HIV patients with breakthrough infection of COVID-19 employed in this study.

## DISCUSSION

Even though the World Health Organization has announced that COVID-19 is no longer a public health emergency of international concern, the genome sequence of SARS-CoV-2 is continually evolving and the new variants (e.g., EG.5.1, JN.1, and KP.2) may have the potential to escape the immunity and threat the people’s health, especially among immunocompromised individuals^27,28^. Therefore, there is still a need for effective COVID-19 vaccines to prevent and control viral transmission. Critically, achieving durable longitudinal immunity — defined as sustained, broad protection against evolving variants over time — remains an unmet challenge. Our data underscore that current vaccine-induced or infection-acquired immunity often wanes rapidly and exhibits limited cross-reactivity against emerging variants. The development and deployment of vaccines for COVID-19, as well as the evaluation of population immunity, continue to be important for public health^29^.

In this work, we developed a high-throughput SARS-CoV-2 bNAb assay to detect a wide range of NAbs to non-Omicron and Omicron variants. Compared to the assay using Luminex technology^10,11^, our SARS-CoV-2 bNAb assay is 35-fold more sensitive, 2.5-fold less costly, and can be accessible to any laboratory that has a standard flow cytometer with the 532 nm and 635 nm lasers. Moreover, the simultaneous detection of NAbs to multiple SARS-CoV-2 variants can be achieved within a single experiment, and the assay can be periodically updated to include new variants. The reliability of NAb detection using our SARS-CoV-2 bNAb assay is demonstrated through comparisons with ELISA-based IgG serology, the cPass sVNT assay, pseudovirus-based neutralization assay and live virus-based neutralization assay (Figure 3).

Using this platform, we constructed a comprehensive landscape of Spike trimer and ACE2 interactions for eleven SARS-CoV-2 variants (Figure 2). The results provide evidence that Omicron BA.1 showed the strongest binding affinity to the ACE2 receptor^30,31^. Notably, the transmission fitness of SARS-CoV-2 variants is governed by multifactorial determinants, including viral binding affinity to host receptors, immune evasion capability against NAbs, intrinsic viral replication efficiency, and host population immunity. These collective factors shape variant-specific epidemic trajectories and underscore the need for continuous surveillance of emerging variants^32^.

Furthermore, we determined the bNAbs in 822 serum samples from human and mouse vaccinated with different SARS-CoV-2 vaccines (WT, D614G, Delta, BA.1, BA.4/5) as well as 890 serum samples from individuals infected with SARS-CoV-2 variants (WT, D614G, Alpha, Delta, BA.1, BA.2, BA.2.76, BA.5.2, BF.7, XBB, and EG.5). The results showed the adaptive changes of population immunity variations induced by vaccination and infection^33,34^.

Importantly, while NAbs serve as critical correlates of protection, emerging evidence highlights the complementary role of T cell effector functions in antiviral immunity. Previous studies, including work from this group evaluating an RBD-nucleocapsid fusion protein as a COVID-19 booster candidate, demonstrate that protein-based vaccines can elicit protective immunity even in the absence of detectable NAbs^35^. CD8+ T cells and CD4+ T cells contribute to viral clearance, memory responses, and protection against variant escape, particularly in immunocompromised populations where antibody responses may be suboptimal. This underscores the value of integrating T cell immunity assessments into vaccine evaluation frameworks.

Finally, we found that in HIV patients with breakthrough infection of COVID-19, individuals with recovered immunity (CD4+ > 500 cells/µL) exhibited significantly higher serum NAb inhibition rates (%) against most of SARS-CoV-2 variants compared to individuals with decreased immunity (CD4+ < 350 cells/µL)^36^. These results demonstrate the significance of our proteomics platform in evaluating the broad protective activity of COVID-19 vaccines and COVID-19 infection in population, which can provide valuable insights for future vaccine development and public health strategies.

The imperative for vaccines inducing longitudinal immunity is further emphasized by our population-level findings. The adaptive changes in humoral immunity we observed across vaccinated and infected cohorts reflect the dynamic interplay between viral evolution and immune erosion. This underscores that narrow-variant targeting in vaccines may offer transient protection at best. To establish persistent population immunity, next-generation vaccines must prioritize conserved epitopes capable of eliciting cross-reactive bNAbs against both current and future variants. Our bNAb platform enables rapid immunogenicity benchmarking against emerging variants (e.g., KP.2), providing essential tools for developing broad-spectrum vaccines that confer variant-resilient longitudinal immunity.

It is important to acknowledge some limitations of our study. First, the *in vitro* detection of NAbs may not represent viral neutralization *in vivo*, although previously reported evidence has shown a good correlation between NAbs detected *in vitro* and *vivo*^8–11^. Second, the relationship between the inhibition levels obtained with the SARS-CoV-2 bNAb assay and the SARS-CoV-2 infection remains unclear. Third, the limited sample size and cohort design constrained our ability to conduct granular sub-analyses, particularly in immunocompromised subgroups (e.g., HIV cohort), to differentiate immune profiles primarily induced by vaccination versus natural infection, and to comprehensively evaluate NAb levels across diverse vaccine types and SARS-CoV-2 variants in larger populations. Fourth, while the assay currently includes 20 SARS-CoV-2 variants, it does not encompass all circulating or emerging lineages (e.g., KP.2). This reflects a practical limitation at the time of study execution. Crucially, the modular design of our bead array platform inherently allows for straightforward extension to incorporate new variants in future iterations. Fifth, the Wellgrow platform utilizes a high-resolution 20-bit analog-to-digital converter (ADC), resulting in raw fluorescence intensity values approximately 100-fold higher than systems using 14-bit ADCs (e.g., Luminex). While this instrumentation difference explains the apparent higher background observed with negative controls (e.g., N protein) and does not reflect reduced specificity, it is an important technical consideration when interpreting raw MFI data. Finally, the SARS-CoV-2 WT was not employed due to the unavailability of this protein during executing experiments. Instead, we employed the D614G variant, which shares high homology with the wild type. Previous studies have demonstrated comparable neutralization potency between the D614G variant and the WT^37^. Additionally, the D614G substitution does not significantly alter SARS-CoV-2 morphology and binding to the ACE2 receptor^38^.

## CONCLUSION

We have successfully developed and validated a high-throughput proteomics platform, enabling the comprehensive evaluation of NAbs against a wide range of SARS-CoV-2 variants. This platform offers an efficient, sensitive, and cost-effective approach for detecting bNAbs within the population, providing an opportunity to guide the design and evaluation of vaccines with enhanced protective efficacy against evolving SARS-CoV-2 variants. Overall, our high-throughput proteomics platform represents a powerful tool for the detection of bNAbs in the population and may inform the development of more effective COVID-19 vaccines and vaccination strategies in the future.

## MATERIALS AND METHODS

### Collection of experimental samples

56 clinical serum samples, collected from healthy individuals at Beijing Ditan Hospital 28 days after administration of the third dose of the COVID-19 vaccine in September 2021 (Table 1). The study received approval from the Beijing Ditan Hospital Ethics Committee (No. 2021-010-01), and all participants provided written informed consent^39^. 35 clinical serum samples were collected from HIV patients with breakthrough infection of COVID-19 at The First Affiliated Hospital of China Medical University from March 1, 2023 to July 1, 2023 (Table 2). By the end of 2022, the predominant variants of the COVID-19 in China were the Omicron BA.5.2 and BF.7. The subjects were HIV patients who were infected with the SARS-CoV-2 during the COVID-19 pandemic in China in 2022^26^. HIV patients the following criteria were included in the study: (1) HIV-1 antibody positive; (2) aged ≥ 18; (3) having COVID-19 infection information; (4) having records of COVID-19 vaccination; (5) being willing to provide blood samples for laboratory testing, such as COVID-19 antibody testing.

A total of 75 clinical serum samples, collected from Peking Union Medical College Hospital (PUMCH) between May and September 2022 and obtained from individuals approximately 180 or 270 days after receiving the third dose of the COVID-19 vaccine, were utilized for correlation analysis with the results from the commercial ELISA-based serology and cPass sVNT assay. The study was approved by the institutional review board of PUMCH (No. K1965-K22C0433), and informed consent was obtained from all participants.

147 clinical serum samples from unvaccinated healthy individuals, along with 135 clinical serum samples from healthy individuals used for correlation analysis with pseudovirus-based neutralization assays (collected 28 days after the second dose of the COVID-19 vaccine), were obtained from Beijing Chaoyang Hospital, in January 2021. The study was approved by the Beijing Chaoyang Hospital Ethics Committee (No. 2020-ke-491), and informed consent was obtained from all participants.

20 serum samples of mouse were collected from 10 mice immunized with WT mRNA vaccines and 10 mice immunized with Omicron BA.1 mRNA vaccines 28 days post-vaccination, obtained under laboratory conditions.

### Preparation of reagents for SARS-CoV-2 bNAb assay

The SARS-CoV-2 bNAb assay was developed in ProteomicsEra Medical Co., Ltd (Beijing, China). Briefly, 3 μg SARS-CoV-2 Spike trimer proteins (Sino Biological., Novoprotein and ACROBiosystems; China) (Table S1) were coupled to 1×10^6^ magnetic-fluorescent beads (Wellgrow Technology Co., Ltd.) as previously described^40^. First, 12 differently encoded magnetic-fluorescent beads, numbered 102-108 and 201-205 (Table S2), were chosen. Then 125 μL containing 1×10^6^ beads were washed with 100 μL ddH2O and activated with 80 μL of “activation buffer” (50 mM 2-(N-morpholino) ethanesulfonic acid (MES) pH 5.0) (Sigma-Aldrich, St. Louis, MO). Then the activated beads were mixed with 10 μL of 50 mg/mL sulfo-N-hydroxysuccinimide (NHS) (Thermo Scientific, USA) and 10 μL of 50 mg/mL 1-Ethyl-3-[3-dimethylaminopropyl]-carbodiimide hydrochloride (EDC) (Solarbio, Beijing, China). After incubation for 20 min at room temperature with gentle mixing, the beads were washed twice with 250 μL “coupling buffer” (50 mM MES pH 5.0) and resuspended in 200 μL coupling buffer. Then 3 μg of Spike trimer proteins were added to the bead solution and incubated for 2 h at room temperature. After washing, the beads underwent blocking with “blocking buffer” (PBST-B, PBS pH 7.4, 0.05% Tween-20, 1% BSA), and the coupled beads were subsequently stored at 2-8 °C in “storage buffer” (PBST-BN, PBS pH 7.4, 0.05% Tween-20, 0.1% BSA, 0.05% NaN3).

### Detection of Spike-ACE2 interactions using SARS-CoV-2 bNAb assay

First, a 50 μL mixed solution containing 12 different SARS-CoV-2 Spike trimer protein-coupled beads (2500 beads per type) (Table S3) was added to each well of a 96-well plate, followed by 50 μL of different concentrations (0, 0.014, 0.041, 0.123, 0.370, 1.111, 3.333, 10 μg/mL) of biotinylated ACE2 (Sino Biological). After incubation for 1 h at room temperature on a shaker, the beads were magnetically separated using a magnetic separator (Wellgrow Technology Co., Ltd.) and washed three times with 100 μL PBST-B (PBS pH7.4, 0.05% Tween-20, 0.1% BSA). After washing, the Spike-ACE2 interaction was detected using 50 μL SA-PE (Thermo Scientific, USA) (2 μg/mL) for 30 minutes at room temperature. Finally, after washing with PBST-B twice, the beads were resuspended in a 200 μL PBST-B solution. The fluorescent signal was then detected at 200 beads/region using the EasyCell flow cytometry (Wellgrow Technology Co., Ltd.) with excitation wavelengths of 532 nm and 635 nm.

### Detection of monoclonal/polyclonal antibodies using SARS-CoV-2 bNAb assay

To detect monoclonal/polyclonal antibodies, a 50 μL mixed solution containing 7 different SARS-CoV-2 Spike trimer protein-coupled beads (2500 beads per type) (Table S4) was added to each well of a 96-well plate. Following that, 50 μL of each antibody solution at different concentrations (0, 0.014, 0.041, 0.123, 0.370, 1.111, 3.333, 10 μg/mL) for anti-Spike antibodies (#26, #20, #21, #22, #23, #73) (Table S5) was added to individual wells of a 96-well plate. After incubation for 2 h at room temperature on a shaker, the beads were washed with PBST-B three times, then the beads were incubated with 50 μL biotinylated ACE2 (0.5 μg/mL) for 1 hour at room temperature. After washing, the binding of ACE2 to Spike trimer proteins were detected using 50 μL SA-PE (2 μg/mL) for 30 minutes at room temperature. Finally, after washing with PBST-B twice, the beads were resuspended in a 200 μL PBST-B solution. The fluorescent signal was then detected at 200 beads/region using the EasyCell flow cytometry (Wellgrow Technology Co., Ltd.) with excitation wavelengths of 532 nm and 635 nm.

### Detection of Serum NAbs using SARS-CoV-2 bNAb assay

First, a 50 μL mixed solution containing 12 different SARS-CoV-2 Spike trimer protein-coupled beads (2500 beads per type) (Table S2) was added to each well of a 96-well plate, followed by 50 μL of the serum samples which were diluted to 1:20 with PBST-B. After incubation for 2 h at room temperature on a shaker, the beads were washed with PBST-B three times. Then the beads were incubated with 50 μL biotinylated ACE2 (0.5 μg/mL) for 1 hour at room temperature. After washing, the binding of ACE2 to Spike trimer proteins were detected using 50 μL SA-PE (2 μg/mL) and incubated the mixture for 30 minutes at room temperature. Finally, after washing with PBST-B twice, the beads were resuspended in a 200 μL PBST-B solution. The fluorescent signal was then detected at 200 beads/region using the EasyCell flow cytometry (Wellgrow Technology Co., Ltd.) with excitation wavelengths of 532 nm and 635 nm.

### Detection of Serum anti-SARS-CoV-2 Spike RBD IgG antibodies using ELISA

The assay was conducted as described previously^20^. A capture sandwich ELISA detection kit (PROPRIUM, Hangzhou, China) was used to detect SARS-CoV-2 antibodies against the Spike protein RBD. The SARS-CoV-2 Spike RBD protein was pre-coated onto the solid phase, allowing it to form an antigen-antibody complex with IgG anti-RBD antibodies from 100 μL of diluted serum samples (1:30 dilution ratio) or standards. After washing, HRP-conjugated anti-human IgG was added to form an antigen-antibody-HRP complex. The substrate solution TMB was then introduced, and the resulting color intensity was proportional to the level of SARS-CoV-2 NAbs. The optical density (OD) at 450 nm was measured, and a threshold of 10 BAU/ml was used to determine sero-positive and negative samples for anti-Spike RBD IgG.

### Detection of Serum NAbs using cPass sVNT assay

The assay was conducted as described previously^20^. Using competitive ELISA, the SARS-CoV-2 sVNT assay (Genscript, Nanjing, China) identified NAbs present in the bloodstream that obstruct the interaction between the viral Spike glycoprotein RBD and human ACE2. Following the manufacturer’s instructions, diluted serum samples (1:10 dilution ratio) and controls were pre-incubated at 37LJ for 15 minutes to allow binding of NAb and HRP-RBD (antigen derived from D614G variant), using a 1:1 volume ratio. Subsequently, the mixture was transferred to a capture plate, pre-coated with hACE2 protein, where NAb-unbound HRP-RBD was captured on the plate, while NAb-bound HRP-RBD remained in the supernatant and was washed away. Next, 100 μL of TMB and 50 μL of stop solution were added, and the plate was read at 450 nm. The sample’s absorbance was inversely correlated with the titers of anti-SARS-CoV-2-NAbs. To ensure result validity, the OD450 values of positive controls (> 1.0) and negative controls (< 0.3) had to fall within specific ranges. An inhibition rate of ≥ 30% was considered positive for SARS-CoV-2 NAb determination. The inhibition rate (%) was calculated as follows: Inhibition rate (%) =LJ(1 - OD value of sample/OD value of negative control)LJ×LJ100%.

### Pseudovirus-based neutralization assay

The SARS-CoV-2 pseudovirus was prepared using the VSV-ΔG system in which the glycoprotein (G) gene was replaced with the firefly luciferase (Fluc) reporter gene^41,42^. The S protein was overexpressed and displayed on the VSV pseudovirus. Following S-ACE2 interaction, the pseudovirus entered the host cell where the Fluc gene was transcribed and translated. The addition of luciferase substrate resulted in luminescence where the amount of luminescence is proportional to the level of pseudoviral entry. The preparation process of SARS-CoV-2 pseudovirus begins with the cloning of the S gene encoding the S protein of the virus. Subsequently, this gene is inserted into a modified VSV backbone, replacing the original G gene. The resulting construct, containing the S protein gene in lieu of the G gene and complemented by the Fluc reporter gene, forms the foundation of the pseudovirus. The pseudovirus equipped with the S protein can interact with the ACE2 receptor on the surface of host cells, thereby facilitating the entry of the pseudovirus into the cells. Once inside the cells, the Fluc gene will be transcribed and translated to produce luciferase.

The assay was conducted following previously described methods^23,43^. Huh7 cells were seeded at a concentration of 2 × 10^4^ cells per well in 96-well plates and incubated until reaching 90-100% confluency, typically around 24 hours. Serum samples, subjected to serial 3-fold dilutions commencing at 1:10, were then incubated with 650 TCID50 of the pseudovirus for a precisely controlled duration of 1 hour at 37 °C. Dulbecco’s modified eagle medium (DMEM) served as the negative control. As a crucial negative control, DMEM was employed. Following the incubation period, the supernatant was carefully aspirated, making way for the addition of luciferase substrate to each well. A subsequent 2-minute incubation in darkness at room temperature facilitated optimal enzymatic reactions. Luciferase activity, indicative of NAb presence, was quantified using the GloMax® 9633 Microplate Luminometer (Promega, Madison, USA).

### Live virus based neutralization assay

The assay was conducted as described previously^43^. To assess the SARS-CoV-2-specific NAb titer in serum, we employed a cytopathic effect (CPE)-based microneutralization assay, utilizing the SARS-CoV-2 virus strain BetaCoV/Beijing/IME-BJ01/2020 (Accession No. GWHACAX01000000) and Vero cells (ATCC, CCL81). Serum samples underwent heat inactivation for 30 minutes at 56 °C and were subsequently two-fold serially diluted (ranging from 1:4 to 1:2048) using DMEM from Thermo Fisher Scientific. These dilutions were then mixed with an equivalent volume of the virus solution to achieve a 50% tissue culture infectious dose (TCID50) of 100 in each well.

The serum-virus mixtures were incubated for 1 hour at 37 °C to allow for sufficient neutralization reactions to occur. Following incubation, the mixtures were gently added to 96-well plates containing semi-confluent Vero cells with a density exceeding 80%. The plates were then incubated for a further 3 days at 37 °C to allow for the development of CPEs on the Vero cells. Using an inverted microscope, the CPEs on the Vero cells were carefully observed and recorded. The neutralizing titer was determined as the reciprocal of the highest sample dilution that successfully protected at least 50% of the cells from CPE. In cases where no neutralization reaction was observable even at the initial serum dilution of 1:4, an arbitrary titer of 2 (half of the limit of quantification) was assigned.

### Data analysis

To compare the inhibition rate (%) of serum NAbs against the D614G variant and other SARS-CoV-2 variants, the raw inhibition rates of different variants were first normalized using minimum-maximum normalization. In order to estimate dose-response relationships, curve fitting and visualization were performed using GraphPad Prism software (version 8.3.0). The Wilcoxon rank sum test was performed using stat_compare_means function implemented in R package ggpubr (version 0.6.1). Inhibition rate (%) of serum NAbs with a p < 0.05 considered as statistically significant. Correlations between the SARS-CoV-2 bNAb assay, ELISA, cPass sVNT assay, pseudovirus-based neutralization assay, and live virus based neutralization assay were determined using Spearman correlation coefficients. Spearman correlation analysis was performed using the stat_cor function implemented in R package ggpubr (version 0.6.1). erum NAbs with a p < 0.05 considered as statistically significant. To assess the reliability of the NAb results obtained from the SARS-CoV-2 bNAb assay, ROC analysis was conducted using roc function implemented in R package pROC (v1.18.2) based on the positive and negative judgment results from ELISA and cPass sVNT assay^44^. Youden index was used in conjunction with the ROC curve to determine the optimal threshold through coords function in pROC (v1.18.2) package. Also, the PPA and NPA between SARS-CoV-2 bNAb assay and cPass sVNT assay were calculated based on the ideal cutoffs from ROC curves. All data analyses were carried out in R (v4.2.3) under the R Studio (v19.1.3) environment.

## Data Availability

All data produced in the present study are available upon reasonable request to the authors

## SUPPORTING INFORMATION

The following is the Supporting Information related to this article: Schematic workflow of the SARS-CoV-2 bNAb assay; Spike trimer-ACE2 interactions across six SARS-CoV-2 VOCs and ACE2 concentrations on different platforms; ACE2 binding to SARS-CoV-2 variants, SARS-CoV, MERS Spike; ROC curves for SARS-CoV-2 bNAb assay, ELISA and cPass sVNT assay; comparison of SARS-CoV-2 bNAb assay to other assay; comparison of NAbs from inactivated vs. recombinant subunit vaccines; serum NAb level difference between vaccinated and unvaccinated groups; lists of: SARS-CoV-2 variant Spike/N proteins, EasyMagPlex beads, coupled proteins/bead codes for Spike-ACE2 & serum NAbs detection, coupled proteins/bead codes for monoclonal/polyclonal NAbs detection, SARS-CoV-2 RBD antibodies, SNR for ACE2 detection on Luminex/Wellgrow (PDF).

## CONTRIBUTIONS

X.Z., and D.H. executed the experiments; M.L., X. Z., X.Z., X.X., T. L., Y. J., and Y.Z. performed the data analysis; H.L., Y.W., X.H., and Q.M. provided the clinical samples. X.Y., Y. L., B.W., X.Z., S.B., S.S., and M.F. drafted, wrote and revised the paper.

## ACKNOWLEDGMENT

This work was supported by the National Key R&D Program of China (2024YFA1307600, 2024YFA1307601, 2022YFE0210400, 2021YFA1301604), Beijing Municipal International Cooperation and Exchange Project (Z231100002723004), Beijing Municipal Natural Science Foundation (M23010, L234034), Capital Health Development Research Project (2024-1-1201 and 2024-1-4093), State Key Laboratory of Proteomics (SKLP-O202205, SKLP-O202007), Guangdong Province Science and Technology Planning Project (2020B1111100006), Guangdong-PHOENIX Center Joint Lab on Chinese Medicine (YN2021DB06), Innovation Team and Talents Cultivation Program of National Administration of Traditional Chinese Medicine (ZYYCXTD-C-202204). We would like to thank the bioinformatics platform of the National Center for Protein Sciences (Beijing) for their support in the data analysis of this project. We also thank Dr. Brianne Petritis for her critical review and editing of this manuscript.

