## Supplementary materials for "A High-Throughput Broad Neutralizing Antibody Assay for Detecting SARS-CoV-2 Variant Immunity in Population"

| **Supporting Information** | **Page** |
| --- | --- |
| Figure S1. Schematic workflow of the SARS-CoV-2 bNAb assay development and validation. | S4 |
| Figure S2. Spike-ACE2 interactions across six SARS-CoV-2 VOCs and ACE2 concentrations on different platforms. | S5-S6 |
| Figure S3. The binding ability of ACE2 to SARS-CoV-2 variant, SARS-CoV and MERS Spike proteins. | S7 |
| Figure S4. ROC curves evaluating the diagnostic performance of SARS-CoV-2 bNAb assay. | S8 |
| Figure S5. Correlation between the inhibition rate (%) of NAbs against the D614G variant and the NAb titers to the D614G variant. | S9 |
| Figure S6. Comparison of NAbs produced between groups receiving the inactivated vaccineor recombinant subunit vaccine. | S10 |
| Figure S7. Difference in serum NAb levels in the vaccination group and un-vaccination group. | S11 |
| Table S1. List of SARS-CoV-2 variant trimer Spike proteins and N protein. | S12 |
| Table S2. List of EasyMagPlex Magnetic-Fluorescent Beads. | S13 |
| Table S3. List of the coupled proteins and corresponding beads codes that were used to detect Spike-ACE2 interactions and serum NAbs. | S14 |
| Table S4. List of the coupled proteins and corresponding beads codes that were used to detect monoclonal/polyclonal NAbs. | S15 |
| Table S5. List of SARS-CoV-2 S protein RBD antibodies. | S16 |
| Table S6. List of the SNR of different concentrations of ACE2 detection using Luminex and Wellgrow platforms. | S17 |

**Contents**

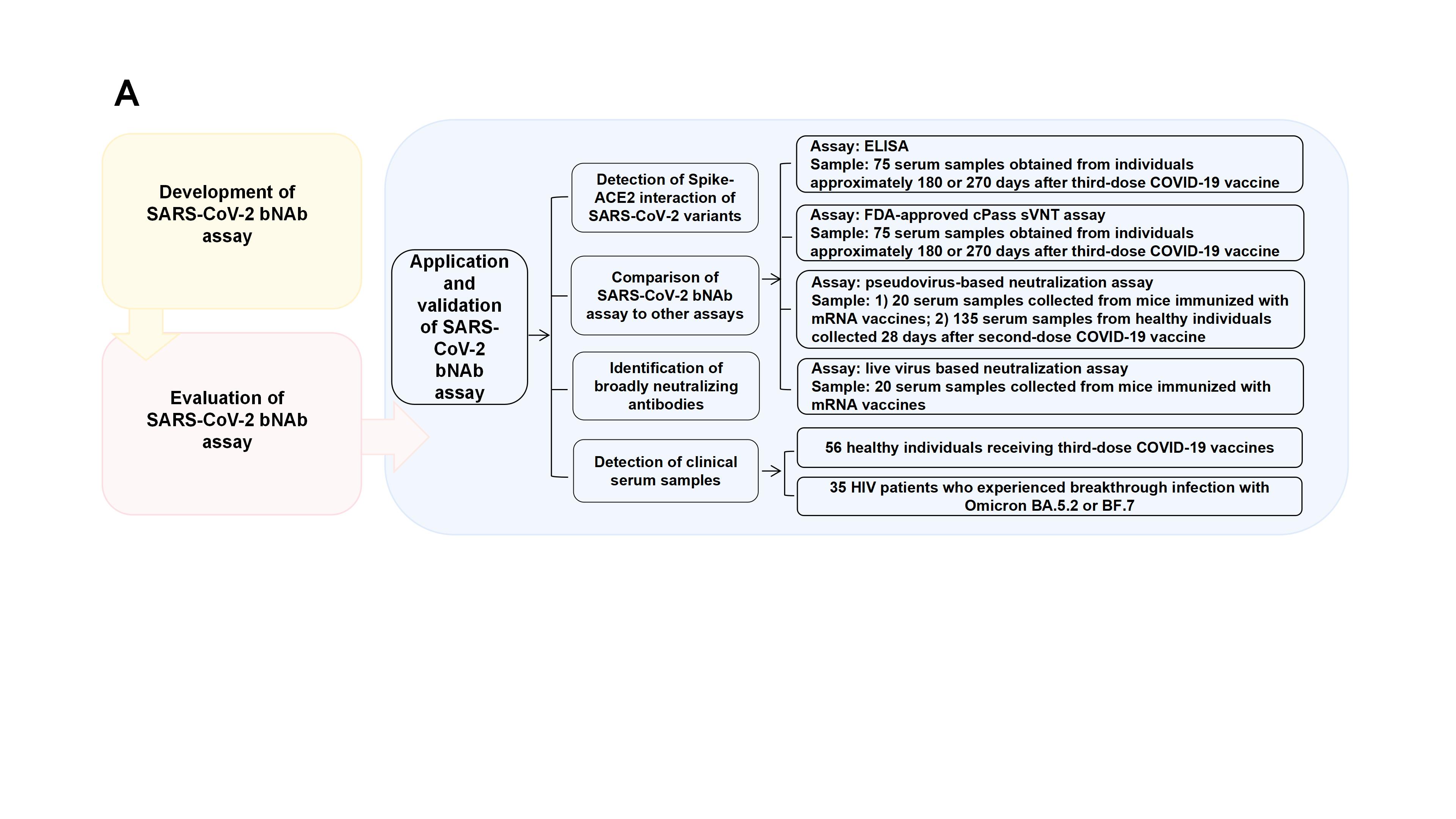

**Figure S1. Schematic workflow of the SARS-CoV-2 bNAb assay development and validation.** (A) Workflow encompassed the development, evaluation, application and validation of the SARS-CoV-2 bNAb assay, with the application and validation phases involving: detection of Spike-ACE2 interaction, comparison to other assays, identification of broadly neutralizing monoclonal/polyclonal antibodies, and detection of NAbs in clinical cohorts. bNAb, broad neutralizing antibody. ACE2, angiotensin-converting enzyme 2. NAb, neutralizing antibody.

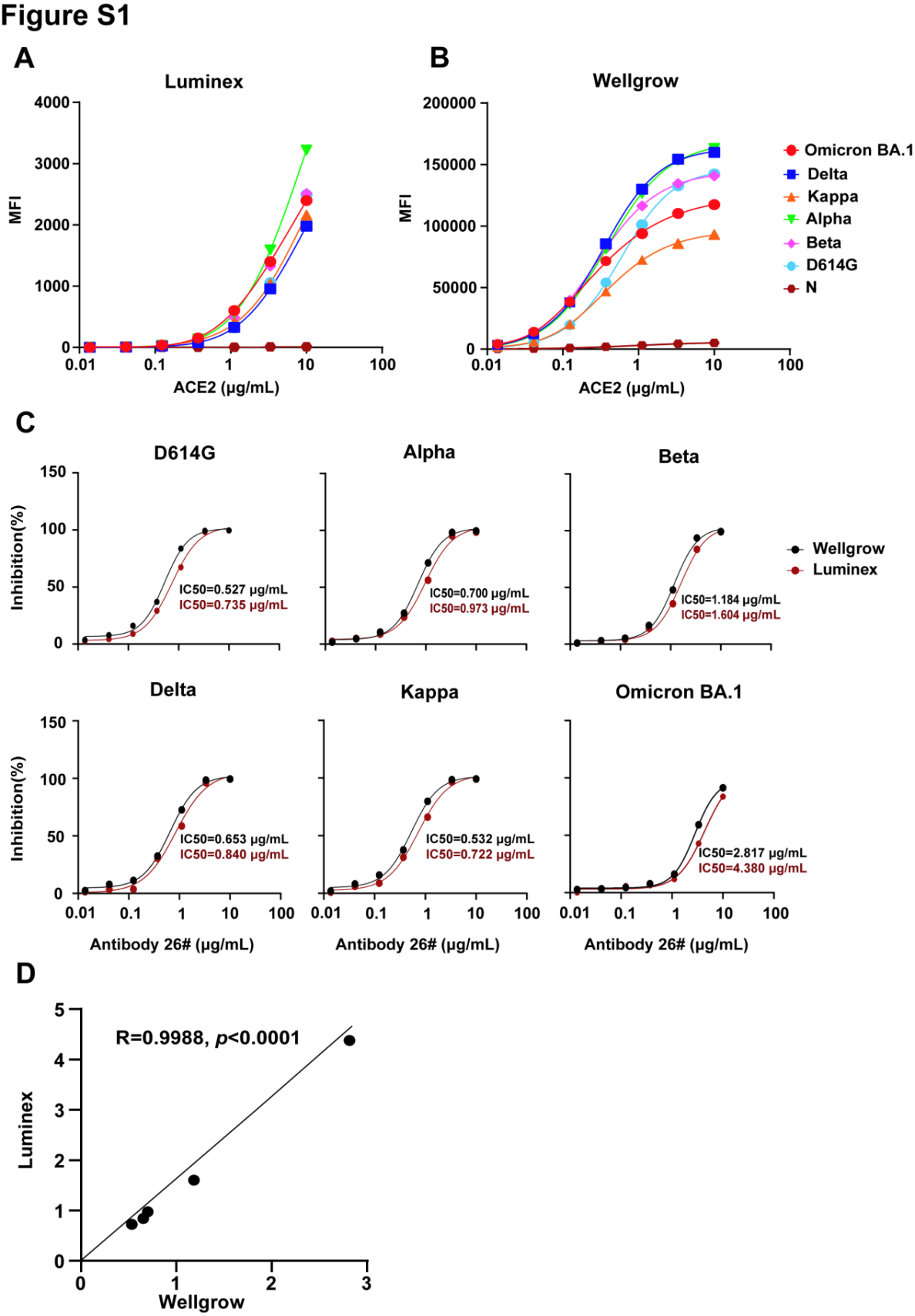

**Figure S2. Spike-ACE2 interactions across six SARS-CoV-2 VOCs and ACE2 concentrations on different platforms.** (A) Dose relationship curve of the interactions between ACE2 and six SARS-CoV-2 VOCs on Luminex platform. (B) Dose relationship curve of the interactions between ACE2 and six SARS-CoV-2 VOCs on Wellgrow platform. The x-axis represents the concentrations of the host ACE2 receptor. The y-axis represents the median of MFI of the SARS-CoV-2 Spike-ACE2 interaction. (C) Dose relationship curve demonstrating the ability of the antibody #26 to inhibit SARS-CoV-2 variant Spike-ACE2 interactions using Wellgrow and Luminex platforms. The x-axis represents the concentrations of the antibody #26. The y-axis represents the inhibition rate (%) of the antibody #26. (D) Correlation analysis of antibody 26# neutralization titers (IC50) using Wellgrow and Luminex platforms. N protein, nucleocapsid protein. ACE2, angiotensin-converting enzyme 2. VOCs, variants of concern. MFI, median fluorescence intensity. IC50, half-maximal inhibitory concentration.

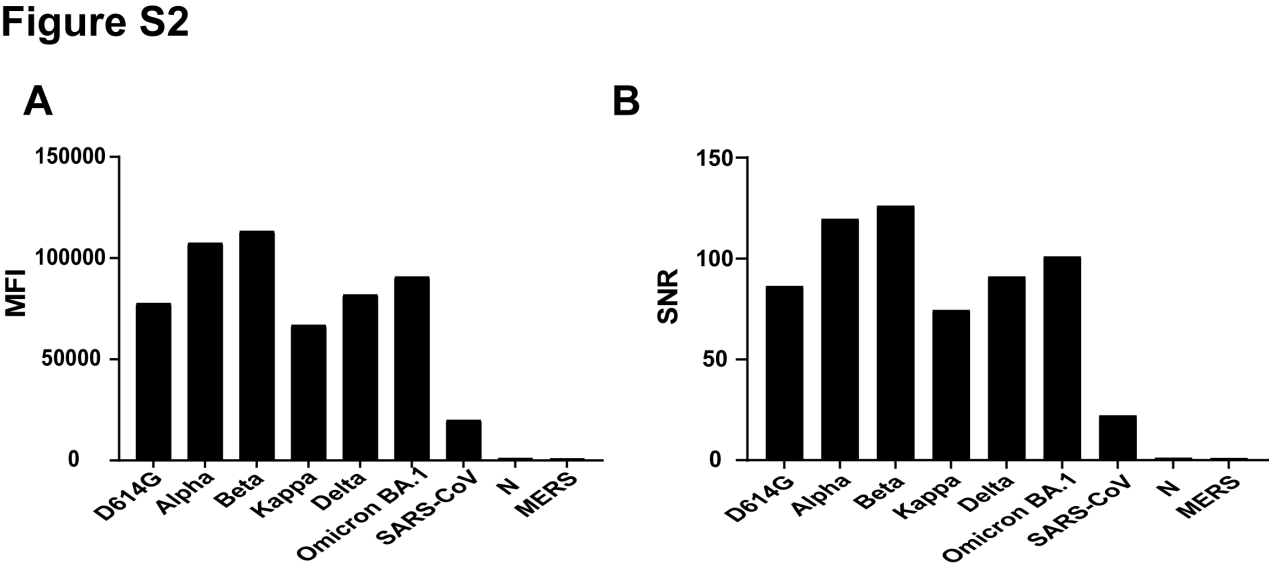

**Figure S3. The binding ability of ACE2 to SARS-CoV-2 variant, SARS-CoV and MERS Spike proteins.** (A) The binding signal of ACE2 to six SARS-CoV-2 variants, SARS-CoV and MERS Spike proteins and the negative control, N protein. (B) The SNR of six SARS-CoV-2 variant, SARS-CoV and MERS Spike proteins, and the negative control N protein as cut off value. ACE2, angiotensin-converting enzyme 2. N protein, nucleocapsid protein. MFI, median fluorescence intensity. SNR, signal to noise ratio.

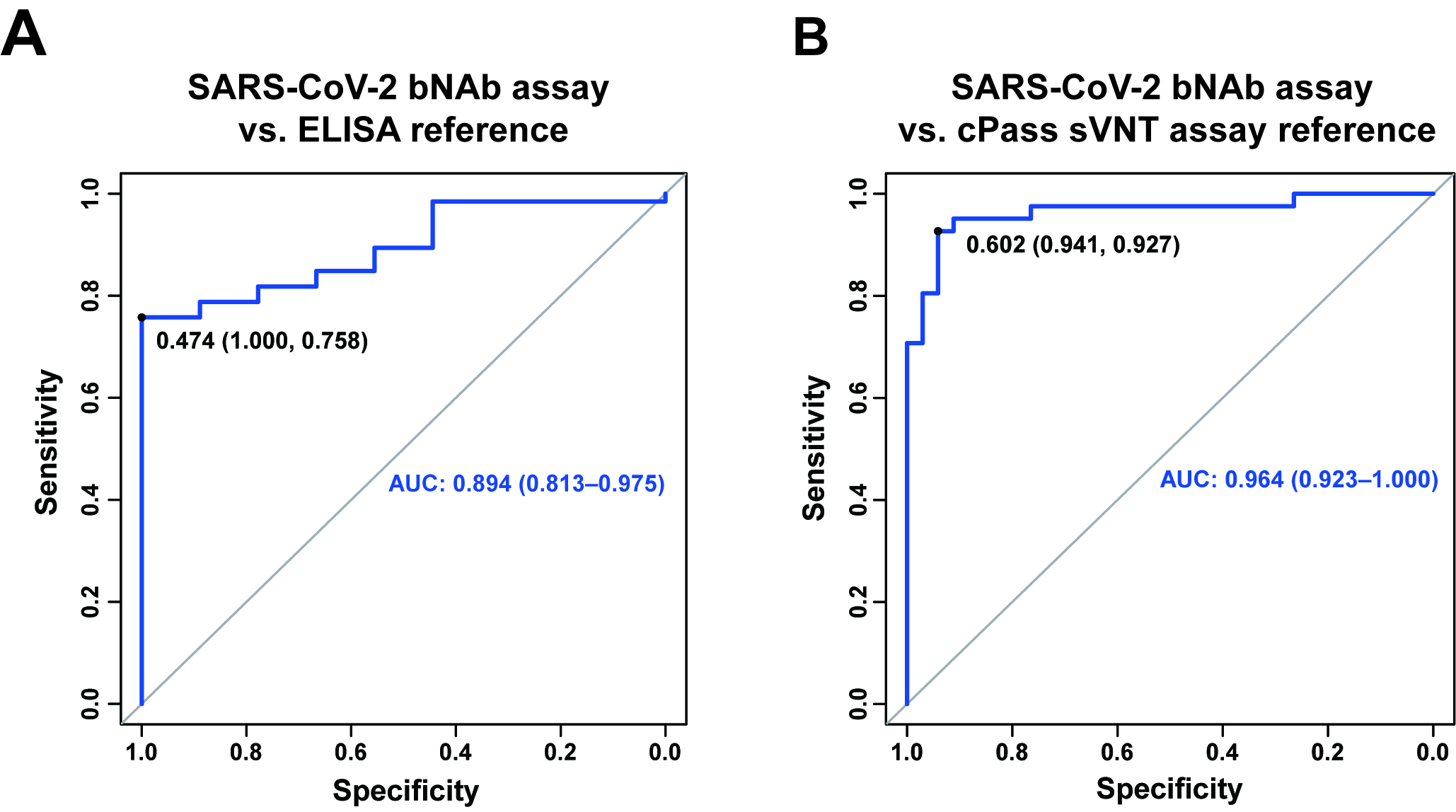

**Figure S4. ROC curves evaluating the diagnostic performance of SARS-CoV-2 bNAb assay.** (A) ROC curve of inhibition rate (%) of NAbs against the D614G variant from the SARS-CoV-2 bNAb assay, using the results of ELISA as reference standard. (B) ROC curve of inhibition rate (%) of NAbs against the D614G variant from the SARS-CoV-2 bNAb assay, using the results of cPass sVNT assay as reference standard. NAb, neutralizing antibody. bNAb, broad neutralizing antibody.

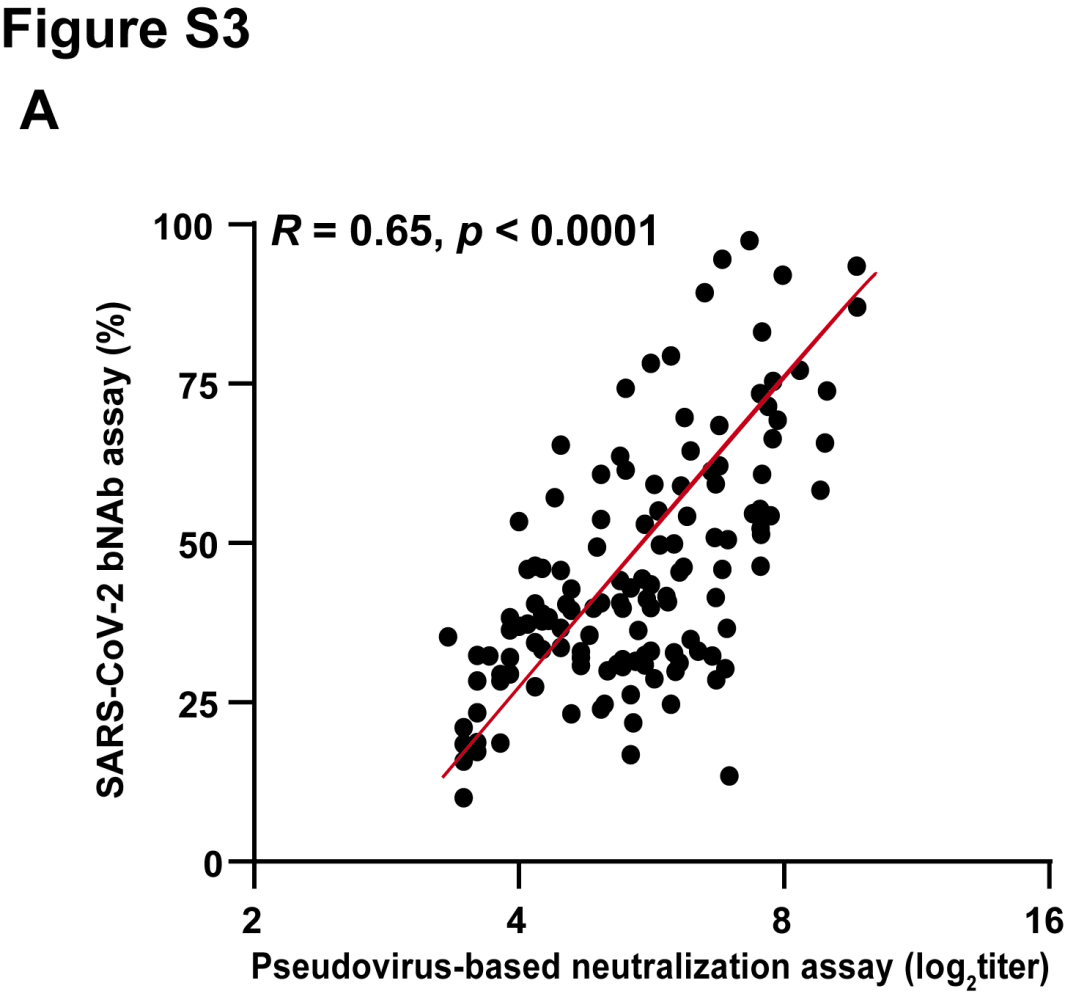

**Figure S5.** **Correlation between the inhibition rate (%) of NAbs against the D614G variant and the NAb titers to the D614G variant.** (A) Correlation between the inhibition rate (%) of NAbs obtained by SARS-CoV-2 bNAb assay and the NAb titers obtained by pseudovirus-based neutralization assay in the serum of 135 individuals with second dose vaccination. NAb, neutralizing antibody. bNAb, broad neutralizing antibody.

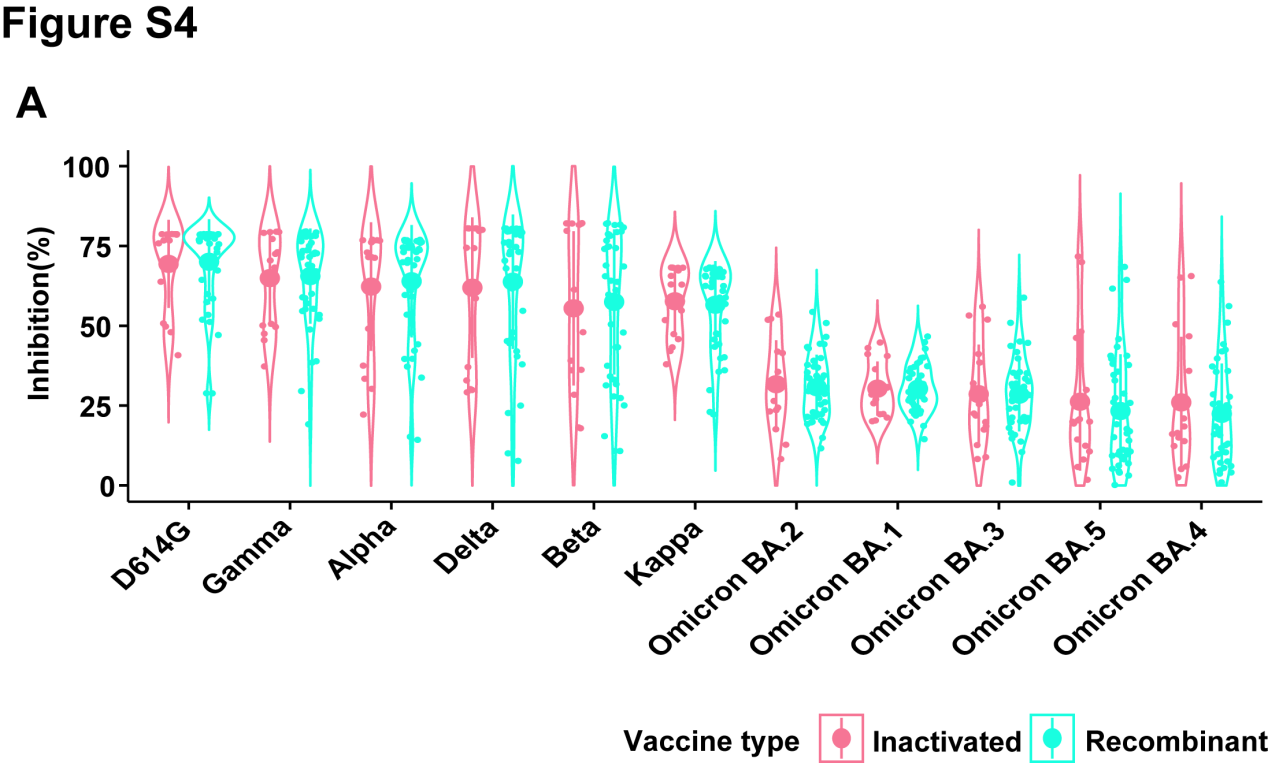

**Figure S6. Comparison of NAbs produced between groups receiving the inactivated vaccine or recombinant subunit vaccine.** (A) Comparison of NAbs produced between groups receiving the inactivated vaccine (Sinovac-CoronaVac) or recombinant subunit vaccine (ZF2001). NAb, neutralizing antibody.

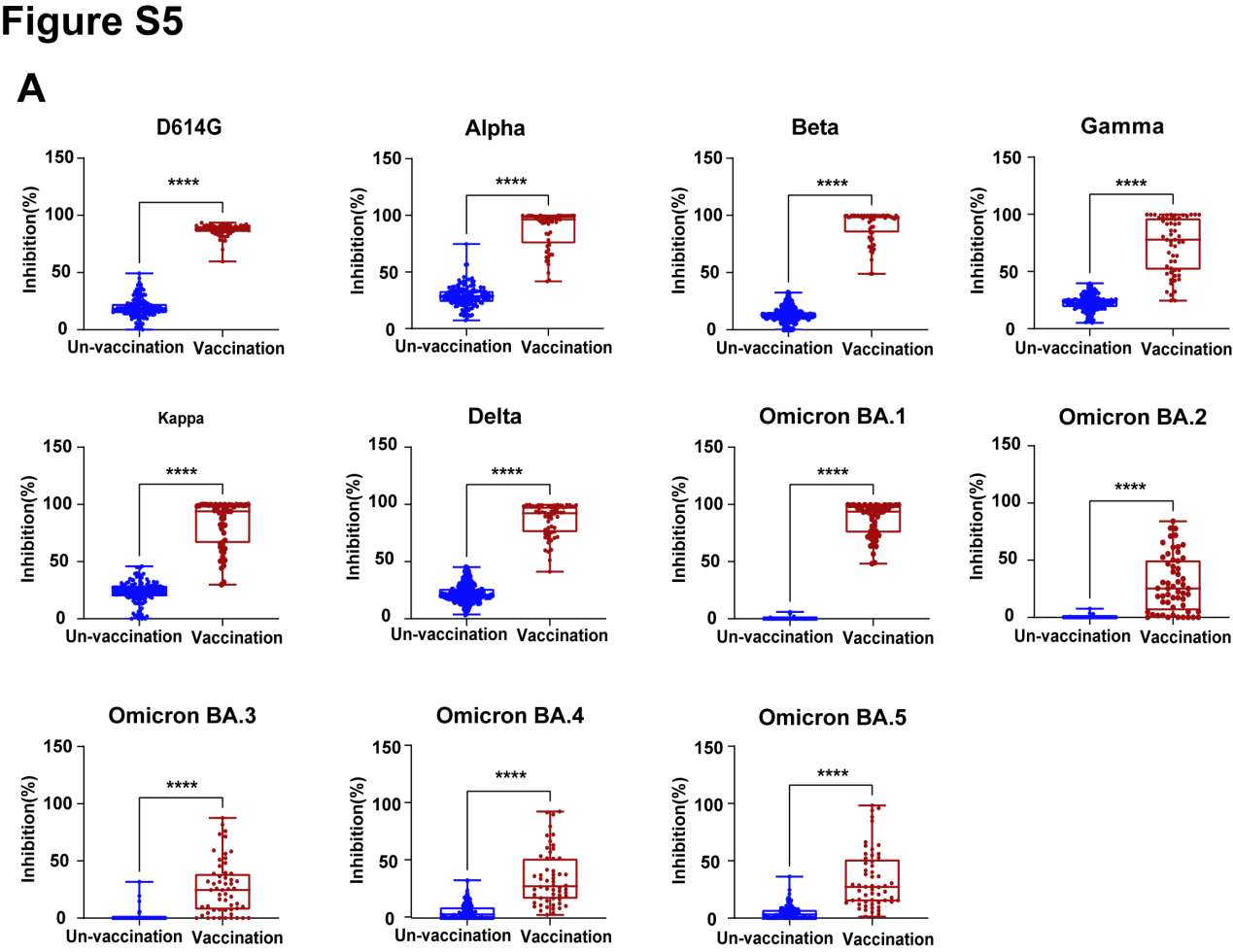

**Figure S7. Difference in serum NAb levels** **in the vaccination group and un-vaccination group.** (A) Inhibition rate(%) of serum NAbs in 11 SARS-CoV-2 variants in the vaccination group and un-vaccination group. Un-vaccination group’s serum samples were obtained from individuals before vaccination (n = 147 ), and vaccination group’s serum samples were collected after three doses of vaccination (n = 56). When the inhibition rate value is negative, it automatically defaults to zero. The serum NAb levels are significantly higher in the vaccination group than un-vaccination group as determined using an Wilcoxon rank sum test. ****, p < 0.0001. NAb, neutralizing antibody.

**Table S1. List of SARS-CoV-2 variant trimer Spike proteins and N protein.**

| **WHO label** | **Product Name** | **Company** | **Catalog No.** |
| --- | --- | --- | --- |
| D614G | SARS-CoV-2 Spike S1+S2 (D614G) trimer Protein (ECD, His tag) | Sino Biological, Inc. | 40589-V08H8 |
| Alpha | SARS-CoV-2 B.1.1.7 Spike S1+S2 trimer Protein (ECD, His tag) | Sino Biological, Inc. | 40589-V08H12 |
| Beta | SARS-CoV-2 B.1.351 Spike S1+S2 trimer Protein (ECD, His tag) | Sino Biological, Inc. | 40589-V08H13 |
| Delta | SARS-CoV-2 B.1.617.2 Spike S1+S2 trimer Protein (ECD, His tag) | Sino Biological, Inc. | 40589-V08H10 |
| Kappa | SARS-CoV-2 B.1.617.1 Spike S1+S2 trimer Protein (ECD, His tag) | Sino Biological, Inc. | 40589-V08H11 |
| Gamma | Recombinant SARS-CoV-2 S-trimer Protein (P.1, C-6His) | Novoprotein | DRA157 |
| Omicron BA.1 | SARS-CoV-2 B.1.1.529 (Omicron) S1+S2 trimer Protein ( ECD, His Tag) | Sino Biological, Inc. | 40589-V08H26 |
| OmicronBA.2 | SARS-CoV-2 (BA.2) Spike S1+S2 trimer Protein (ECD, His Tag) | Sino Biological, Inc. | 40589-V08H28 |
| OmicronBA.3 | SARS-CoV-2 Spike Trimer, His Tag (BA.3/Omicron) | ACROBiosystems | SPN-C5225 |
| OmicronBA.4 | SARS-CoV-2 Spike Trimer, His Tag (BA.4/Omicron) | ACROBiosystems | SPN-C5229 |
| OmicronBA.5 | SARS-CoV-2 Spike Trimer, His Tag (BA.5/Omicron) | ACROBiosystems | SPN-C522e |
| N protein | SARS-CoV-2 (2019-nCoV) Nucleocapsid-His recombinant Protein | Sino Biological, Inc. | 40588-V08B |

**Table S2. List of EasyMagPlex Magnetic-Fluorescent Beads.**

| **Beads Code** | **Product Name** | **Company** | **Catalog No.** |
| --- | --- | --- | --- |
| 102 | EasyMagPlex Magnetic-Fluorescent Beads | Wellgrow Technology | CM020102 |
| 103 | EasyMagPlex Magnetic-Fluorescent Beads | Wellgrow Technology | CM020103 |
| 104 | EasyMagPlex Magnetic-Fluorescent Beads | Wellgrow Technology | CM020104 |
| 105 | EasyMagPlex Magnetic-Fluorescent Beads | Wellgrow Technology | CM020105 |
| 106 | EasyMagPlex Magnetic-Fluorescent Beads | Wellgrow Technology | CM020106 |
| 107 | EasyMagPlex Magnetic-Fluorescent Beads | Wellgrow Technology | CM020107 |
| 108 | EasyMagPlex Magnetic-Fluorescent Beads | Wellgrow Technology | CM020108 |
| 201 | EasyMagPlex Magnetic-Fluorescent Beads | Wellgrow Technology | CM020201 |
| 202 | EasyMagPlex Magnetic-Fluorescent Beads | Wellgrow Technology | CM020202 |
| 203 | EasyMagPlex Magnetic-Fluorescent Beads | Wellgrow Technology | CM020203 |
| 204 | EasyMagPlex Magnetic-Fluorescent Beads | Wellgrow Technology | CM020204 |
| 205 | EasyMagPlex Magnetic-Fluorescent Beads | Wellgrow Technology | CM020205 |

**Table S3. List of the coupled proteins and corresponding beads codes that were used to detect Spike-ACE2 interactions and serum NAbs.**

| **Protein** | D614G | Alpha | Beta | Gamma | Delta | Kappa |
| --- | --- | --- | --- | --- | --- | --- |
| **Beads Code** | 102 | 204 | 103 | 104 | 105 | 106 |
| **Protein** | Omicron BA.1 | Omicron BA.2 | Omicron BA.3 | Omicron BA.4 | Omicron BA.5 | N protein |
| **Beads Code** | 107 | 108 | 202 | 203 | 205 | 201 |

**Table S4. List of the coupled proteins and corresponding beads codes that were used to detect monoclonal/polyclonal NAbs.**

| **Protein** | D614G | Alpha | Beta | Delta | Kappa | Omicron BA.1 | N |
| --- | --- | --- | --- | --- | --- | --- | --- |
| **Beads Code** | 102 | 204 | 103 | 105 | 106 | 107 | 201 |

**Table S5. List of SARS-CoV-2 S protein RBD antibodies.**

| **ID** | **Company** | **Catalog No.** | **Antigen** | **MAb/PAb^*^** |
| --- | --- | --- | --- | --- |
| #20 | Sino Biological, Inc. | 40150-D001 | SARS-CoV S protein RBD | MAb |
| #21 | Sino Biological, Inc. | 40150-D002 | SARS-CoV S protein RBD | MAb |
| #22 | Sino Biological, Inc. | 40150-D003 | SARS-CoV S protein RBD | MAb |
| #23 | Sino Biological, Inc. | 40150-D004 | SARS-CoV S protein RBD | MAb |
| #26 | Sino Biological, Inc. | 40592-T62 | SARS-CoV S protein RBD | PAb |
| #73 | Bioworld Technology, Co. | HMB002 | SARS-CoV S protein RBD | MAb |

**Table S6. List of the signal to noise ratio (SNR) of different concentrations of ACE2 detection using Luminex and Wellgrow platforms.**

|  | **SNR** | | | | | | | | | | | | | |
| --- | --- | --- | --- | --- | --- | --- | --- | --- | --- | --- | --- | --- | --- | --- |
|  | **D614G** | | **Alpha** | | **Beta** | | **Delta** | | **Kappa** | | **Omicorn BA.1** | | **N protein** | |
| **ACE2**  **(μg/mL)** | **W*** | **L*** | **W** | **L** | **W** | **L** | **W** | **L** | **W** | **L** | **W** | **L** | **W** | **L** |
| **10.000** | 1475.51 | 107.12 | 1896.44 | 144.22 | 1915.00 | 140.57 | 1756.90 | 128.65 | 1058.54 | 109.00 | 1269.25 | 97.82 | 32.91 | 1.63 |
| **3.333** | 1375.75 | 45.95 | 1783.64 | 71.70 | 1830.20 | 74.65 | 1695.82 | 62.68 | 975.26 | 53.10 | 1192.15 | 57.54 | 14.83 | 1.44 |
| **1.111** | 1048.04 | 14.47 | 1471.18 | 25.76 | 1583.45 | 28.19 | 1427.86 | 22.02 | 824.66 | 21.28 | 1015.33 | 25.32 | 5.97 | 1.11 |
| **0.370** | 561.60 | 4.23 | 947.25 | 7.16 | 1111.94 | 7.69 | 940.63 | 5.94 | 533.31 | 6.05 | 774.11 | 7.19 | 2.84 | 1.20 |
| **0.123** | 202.57 | 1.77 | 420.46 | 2.49 | 547.71 | 2.68 | 416.90 | 2.34 | 230.55 | 2.18 | 415.74 | 2.57 | 1.61 | 1.06 |
| **0.041** | 56.88 | 1.21 | 132.79 | 1.41 | 184.33 | 1.31 | 132.80 | 1.34 | 71.03 | 1.23 | 151.67 | 1.37 | 1.19 | 1.10 |
| **0.014** | 16.02 | 0.98 | 35.91 | 1.17 | 54.05 | 1.07 | 37.98 | 1.03 | 21.42 | 1.10 | 45.10 | 1.07 | 1.06 | 1.03 |
| **Mean** | 676.62 | 25.10 | 955.38 | 36.27 | 1032.38 | 36.60 | 915.56 | 32.00 | 530.68 | 27.70 | 694.76 | 27.55 | 8.63 | 1.22 |
| **Fold Change**  **(W/L)** | 26.95 | | 26.34 | | 28.21 | | 28.61 | | 19.16 | | 25.21 | | 7.05 | |
| **Mean of**  **Fold Change** | 23.08 | | | | | | | | | | | | | |
| *W and L are abbreviations for Wellgrow and Luminex, respectively | | | | | | | | | | | | | | |
